# Depression in Premanifest Huntington’s Disease: Aberrant Effective Connectivity of Striatum and Default Mode Network

**DOI:** 10.1101/2024.03.17.24304066

**Authors:** Tamrin Barta, Leonardo Novelli, Nellie Georgiou-Karistianis, Julie Stout, Samantha M Loi, Yifat Glikmann-Johnston, Adeel Razi

**Affiliations:** Turner Institute of Brain and Mental Health at the School of Psychological Sciences, and, Faculty of Medicine, Nursing and Health Sciences, Monash University; Department of Psychiatry, University of Melbourne; Neuropsychiatry Centre, Royal Melbourne Hospital, Parkville, Australia; Monash Biomedical Imaging, Monash University, Clayton, Victoria, Australia; Wellcome Centre for Human Neuroimaging, University College London, London, United Kingdom; CIFAR Azrieli Global Scholars Program, CIFAR, Toronto, Ontario, Canada

## Abstract

2.

**Background:** Depression frequently precedes motor symptoms in Huntington’s disease gene expansion carriers (HDGECs), yet the neural mechanisms remain poorly characterized.

**Objectives:** We investigated effective connectivity between the default mode network and striatal regions in HDGECs.

**Methods:** We analyzed 3T resting-state functional magnetic resonance imaging data from 98 HDGECs (48.98% females; mean age = 42.82 years). Spectral dynamic causal modeling estimated subject-level connectivity, while parametric empirical Bayes determined group-level effective connectivity differences between participants with a diagnosed depression history and those without, across current, remitted, and never-depressed states. Brain-behavior associations with clinical depression measures were examined.

**Results:** Model estimation was excellent (89.82% variance-explained). HDGECs with depression history showed decreased inhibitory posterior cingulate cortex -to-hippocampal connectivity, increased hippocampus-to-posterior cingulate cortex inhibition, and increased inhibitory influence of striatum on default mode network. HDGECs with a depression history showed increased inhibitory striatal influence on DMN, including left putamen, a propensity for right hippocampal involvement, and disinhibitory posterior cingulate-hippocampal connectivity. Current versus never-depressed comparisons revealed more pronounced dysconnectivity, with stronger striatum-to-network connections. Current versus remitted depression exhibited distinct patterns with increased medial prefrontal cortex-to-posterior cingulate cortex connectivity, increased medial prefrontal cortex self-connectivity and decreased posterior cingulate cortex-to-medial prefrontal cortex connectivity.

**Conclusions:** These findings establish distinct striatal-network interaction patterns in depression for HDGECs that differ from non-neurological depression. Our findings suggested posterior DMN—posterior cingulate and hippocampus—as drivers of depression for HDGECs, and potential compensation of right DMN in keeping with compensatory patterns broadly in HD. These connectivity patterns could serve as functional biomarkers for depression in HDGECs.

## 3. Introduction

Depression is common for premanifest Huntington’s disease (HD) gene-expansion carriers (HDGECs), many years before the onset of manifest (clinical) diagnosis (1,2). Up to 20% of HDGECs experience depression (3), at rates higher than the general population (4), and with significant symptom increases in the lead up to manifest diagnosis (3). This elevated prevalence carries clinical importance, as depression in HDGECs is associated with increased suicidal behavior (5), and cognitive impairment (6,7). Preclinical evidence has implicated huntingtin (the protein impacted by the gene-expansion) in depression, for example, depressive behaviors in HD mice can be prevented through modulating huntingtin (8). Despite well-characterized early cortico-striatal degeneration (9), the pathophysiology of depression in HDGECs remains poorly understood.

The etiology of depression in HDGECs is complex, involving psychosocial and neuropathological bases (10). Some people with HD report depression despite previous resilience and support a neurobiological underpinning (10). HDGECs report elevated depressive symptoms and depressive disorders compared to the general population and at-risk relatives (1,2,11). Further, HDGECs have significantly higher depressive symptoms than at-risk relatives even when blinded to their genetic status (3). These findings demonstrate disease-specific progression for depression in premanifest HDGECs and suggest a neurobiological basis.

HD is characterized by early and severe striatal atrophy (i.e., caudate and putamen) including in HDGECs (12–14). Beyond the striatum in premanifest HD, functional connectivity changes, including occipital and frontal hyperconnectivity, are shown over 20 years before manifest diagnosis (15). Functional brain hyperconnectivity transforms to hypoconnectivity as disease burden grows and HD progresses (15,16). Additionally, there is increased effective connectivity of interhemispheric anterior default mode network (DMN) regions for premanifest HDGECs (16). The DMN is a very well-described task-negative (active during rest) brain network, comprising anterior—medial prefrontal cortex (MPFC)—and posterior—posterior cingulate cortex (PCC)—hubs (17). Recent research has proposed the PCC as a disease epicenter for HDGECs, with significant decreased structural connectivity prior to manifest diagnosis (15). In sum, striatal and DMN changes appear as early neural signatures of HD, preceding manifest diagnosis.

DMN and striatal dysconnectivity provides a potential mechanism for depression in HDGECs, as it is a neural marker of major depression (18,19). In non-neurodegenerated populations, there is an evidence linking depression to white and grey matter changes in DMN and striatum (20), aberrant striato-frontal network topology (21), increased functional connectivity between caudate and prefrontal cortices (22) and within DMN (MPFC, PCC, and hippocampus) including the caudate (23), as well as increased effective connectivity from MPFC to posterior DMN (24). Collectively, these findings demonstrate depression as a disorder of striatal and DMN network dysfunction.

Dysconnection of large-scale, functional networks has been proposed as the neuropathological basis for depression in HD (25). For HDGECs, depressive symptoms associate with reduced cortical thickness in DMN and striatum across premanifest HDGECs and manifest HD (26), decreased structural connectivity between anterior DMN and caudate (27), and increased functional connectivity from anterior to posterior DMN (27). The hippocampus, considered a DMN subsystem (28), shows altered function linked to depressive behaviors in preclinical HD models during early disease stages (29,30). Both structural and functional hippocampal connectivity changes have been associated with depression for HDGECs (27). Despite this growing evidence base, directed (effective) connectivity between DMN and striatal dysconnectivity in HD-related depression remains understudied, motivating our focus on causal interactions in subsequent analyses.

Existing studies of depression in HD rely on functional connectivity (18,23,27,28), a correlation-based measure reflecting undirected dependencies between regions (31,32). Effective connectivity allows for the investigation of complex brain dynamics as a model-based, biophysically informed measure that infers the directed influence of a neural system over another, including both valence (inhibition and excitation) and strength (increasing or decreasing) of connectivity (33). Both functional and effectivity connectivity are calculated from the blood-oxygen-level-dependent (BOLD) signal, which is comprised of neural, hemodynamic—such as cerebral blood flow—and noise components. The neurodegenerative process in HD impacts the BOLD signal with severe and early cerebral hypoperfusion in frontotemporal and cingulate regions (34). While functional connectivity analyses may erroneously identify signal differences due to CBF reductions as neuronal differences (35), effective connectivity overcomes this potential confound by modeling relative contributions of neuronal and neurovascular mechanisms. We applied spectral dynamic causal modeling (spectral DCM), a popular method for inferring effective connectivity using resting-state fMRI data. DCM uses Bayesian model fitting and discriminates inhibition and excitation of both bottom-up and top-down connections, allowing for inference of the direction and strength of connectivity between brain regions and within region self-connectivity (i.e., synaptic activity)(33). Further, the framework measures the association between changes in effective connectivity and behavioral measures (36).

We investigated whether DMN and striatal effective connectivity changes underpin depression for premanifest HDGECs. We hypothesize increased effective connectivity from MPFC to PCC and hippocampus for HDGECs with a history of depression compared to those without. We further hypothesize decreased effective connectivity from MPFC to putamen and caudate for HDGECs with a history of depression compared to those without, reflecting early and severe degeneration of these regions. We expect to see a positive association between current depressive symptoms and increased connectivity from anterior to posterior DMN.

## 4. Methods and Material

### 4.1. Participants

This study used Track-On HD data, with full protocol previously reported (14,37). Track-On HD was approved by local ethics committees at participating sites and participants provided written informed consent in accordance with the Declaration of Helsinki. The study included clinical and imaging data from 98 HDGECs (48 female, M_age_ = 45.66, *SD* = 9.95). All participants had a cytosine, adenine, guanine (CAG) repeat length greater than 39. Exclusion criteria included history of significant head injury and major neurological conditions, excluding HD (14,37). HDGECs were not excluded based on medication use, unless part of a therapeutic trial. Comorbid medical conditions, including depression, and associated International Statistical Classification of Diseases and Related Health Problems 10th Revision (ICD-10) classifications were collected and verified by trained clinicians. Using this data, participants were categorized into two groups based on history of depression (current and remitted) or no history of diagnosed depression. ICD-10 classifications and episode details are reported in eTable 1.

#### 4.1.1. Clinical Measures

Track-On HD involved a comprehensive neuropsychiatric battery, including demographic and clinical measures. The study controlled for concomitant regular (daily) mood medication use, including antipsychotics, benzodiazepines, selective serotonin reuptake inhibitors (SSRIs), and non-SSRIs. Medications were used commonly for depression, but also anxiety, post-traumatic symptoms, insomnia, and irritability. A total of 25 participants were taking medications, with 23 (92%) taking only one. Consistent with existing HD neuroimaging studies using the same cohort (14,27,37–42), we included a covariate to account for medication use. Medication type, regime, and indication are reported in eTable 2.

Four HD-related measures were included. The UHDRS Total Motor Score (TMS) is a 15-item clinician-rated severity of motor signs with a maximum score of 124 (43). The CAG-Age Product (CAP) score models the interaction of age and CAG repeat length on disease progression where the score is 100 at expected age of diagnosis (44–47). The Disease Burden Score (DBS) indexes exposure to huntingtin by modelling age and CAG repeat, and includes a constant of 35.5 (46). The HD Integrated Staging System (HD-ISS) incorporates pathophysiology alongside clinical and functional changes, and includes stages 0–3 based on genetic confirmation, striatal volume changes, cognitive impairment, and functional decline (48).

#### 4.1.2. Mood Measures

The study included two self-report measures of depressive symptoms: the Beck Depression Inventory, 2nd Edition (BDI-II)(49) and Hospital Anxiety and Depression Scale, Depression Subscale (HADS-D; (50). The BDI-II has 21 items on a 4-point Likert scale (maximum score: 63) assessing depressive symptoms over a two-week period (49). The BDI-II has been recommended in HD (51) and a cut-off score of 10/11 discriminates clinically elevated depression in HD, with sensitivity of 1.00 and specificity of 0.66 (52). The HADS-D has seven items on a 4-point Likert scale (maximum score: 21) assessing depressive symptoms over one week (50). Research in HD suggests a cut-off of 6/7 discriminates clinically elevated depression, with sensitivity of 1.00, and specificity of 0.82 (52). These cut-offs were used to classify clinically meaningful depressive symptoms.

#### 4.1.3. Demographic Analyses

Demographic variables were compared using Pearson’s chi-squared test or linear model ANOVA. Assumptions were met (see supplementary results). Some variables demonstrated non-normal distributions: this was considered acceptable as it did not impact spectral DCM analyses.

### 4.2. Statistical Analyses

See Fig. 1 for an outline of the statistical pipeline, described in detail below.

**Fig. 1:**
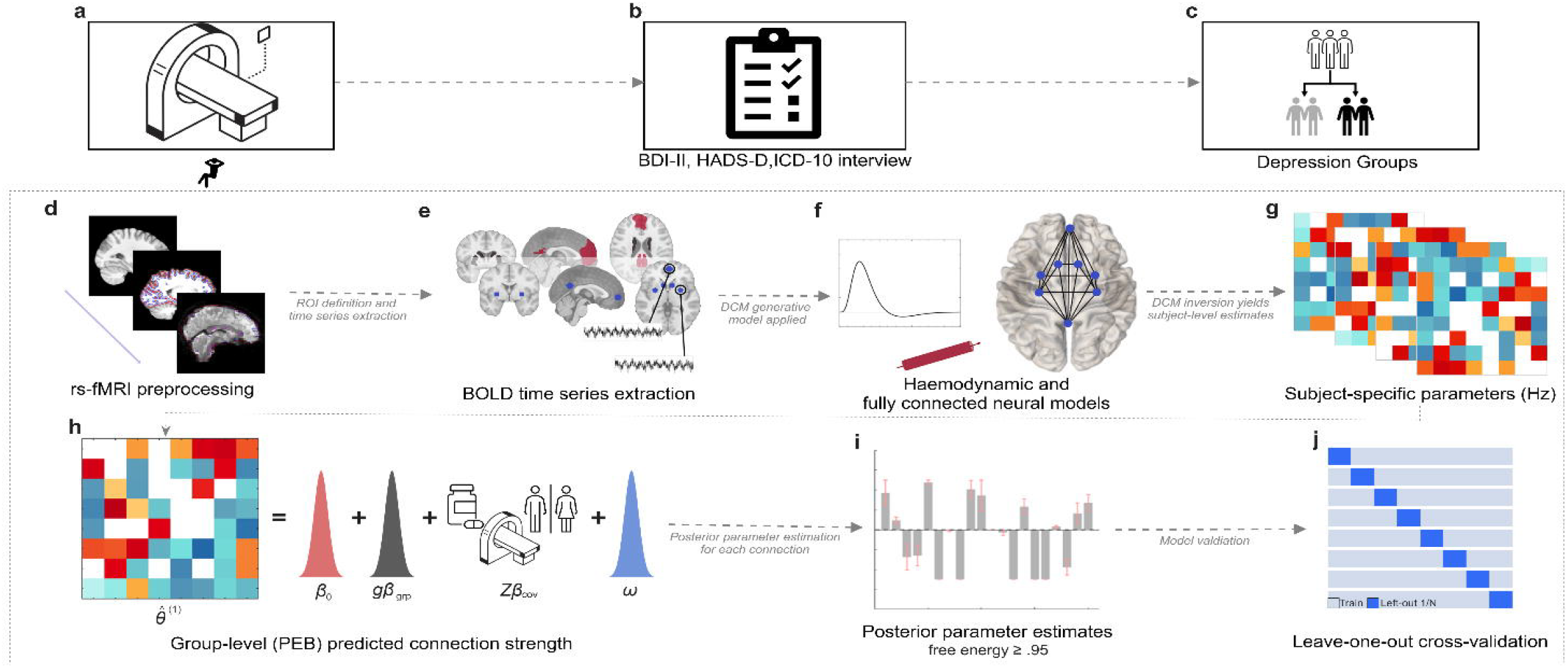
Statistical Analysis pipeline. **(a-c)** Study protocol. **(a)** 3T resting-state functional MRI acquisition. **(b)** Clinical assessment comprised International Statistical Classification of Diseases and Related Health Problems 10th Revision (ICD-10) classifications for depression history categorization and symptom severity measures including Beck Depression Inventory, 2nd Edition (BDI-II) and Hospital Anxiety and Depression Scale, Depression Subscale (HADS-D). **(c)** Participant grouping based on depression history status (HDGECs with depression history group vs. HDGECs without depression history group). **(d–j)** Spectral dynamic causal modeling analysis pipeline. **(d)** pre-processing pipeline using fMRIPrep v21.02.2 and MRIQC v22.0.6, with quality assessment and artifact removal. **(e)** BOLD time series extraction from eight regions of interest, defined as either 6-or 8-mm spheres centered on MNI coordinates and constrained by neuroanatomical masks, with representative time series extraction shown. **(f)** Spectral dynamic causal modeling implementation using a hierarchical generative model incorporating both hemodynamic and (fully connected) effective connectivity parameters, where the Bayesian model fitting procedures optimizes these model parameters to best explain observed cross-spectral densities. **(g)** Subject-specific connectivity parameter estimation yielding 8×8 effective connectivity matrices, with color-coded values representing connection strength and directionality (Hz) between region pairs. **(h)** Shows the predicted connection strength for each region-to-region effect (in Hertz), where θ_i_^(1)^ is participant *i*’s posterior estimate of the connection, β_0_ is the group mean at mean-centered covariates, gβ_grp_ is the group difference term, Zβ_cov_ represents covariate effects (sex, medication, and scanner differences), and ω is the parameter-level residual. **(i)** Posterior parameter estimates, with posterior probability ≥.95, following Bayesian model reduction to identify connections showing reliable group differences. **(j)** Leave-one-out cross-validation procedure excluded one participant iteratively to assess model generalizability and out-of-sample prediction accuracy.

#### 4.2.1. Data Acquisition and Pre-processing

3T MRI data were acquired on two scanner systems at four sites: Philips Achieva (Vancouver and Leiden) and Siemens TIM Trio (London and Paris). Pre-processing was performed using fMRIPrep v21.02.2 (53,54) and MRIQC v22.0.6 (55), using FreeSurfer v-6.0.1 (56). In brief, the pipeline included slice-timing correction, realignment, spatial normalization to Montreal Neurological Institute (MNI) space, and spatial smoothing by a 6 mm full-width half-maximum Gaussian kernel. See supplementary methods for acquisition parameters and preprocessing pipeline.

#### 4.2.2. Selection and Extraction of Volumes of Interest

The selection of regions of interest (ROIs) was based on previous research (17,57) and Neurosynth (58). ROIs and MNI coordinates were MPFC [3,54,-2], PCC [0,-52,26], hippocampus (left [-29,-18,-16], right [29,-18,-16]), caudate (left [-10,14,0], right [10,14,0]), and putamen (left [-28,2,0], right [-28,2,0]). Each ROI time series was calculated as the first principal component of the voxels’ activity within an 8 mm sphere for MPFC and PCC and 6 mm sphere for all other regions and further constrained within anatomical mask boundaries. Masks were chosen using Stanford Willard Atlas(59) for MPFC and WFU PickAtlas (60) for all other regions. Preprocessed data underwent smoothing and a generalized linear model regressed white matter and cerebrospinal fluid signals and 6 head motion parameters (3 translation and 3 rotational).

#### 4.2.3. Dynamic Causal Modelling

Effective connectivity was estimated using spectral DCM. Spectral DCM employs a linear state-space model to fit the cross-spectral densities of the observed BOLD signal. These data features are the frequency domain equivalent of cross-correlation functions and provide computational efficiency when inverting large-scale brain networks (61).

A fully connected within-subject DCM model with no exogenous inputs (i.e., resting-state) was specified using the ROIs. After specification, the model was inverted by fitting the DCM forward model (i.e., its parameters) to provide the best prediction for observed cross-spectral densities. For each subject, spectral DCM estimated the effective connectivity strength between each pair of ROIs and corresponding uncertainty.

The inferred effective connectivity from the first-level analysis was used for hypothesis testing of between-subjects’ effects. Parametric empirical Bayes estimated the effect of a previous history of depression on each connection (36). The parametric empirical Bayes procedure modeled within-subject connectivity parameters, including the expected strength, parameter covariance, and unexplained (between-subject) noise to estimate unique effects of variable of interest (depression history, depression symptoms) on connectivity parameters, controlling for model covariates. Covariates included mood medication use, sex, and the first principal component of between scanner effects (eFig. 8). Lastly, Bayesian model reduction was employed as an efficient form of Bayesian model selection (36). Only connections exceeding the threshold of free energy ≥ 0.95 (strong evidence) are reported. Self-connections in DCM are modulators controlling the balance between excitation and inhibition, and are neurobiologically explained as modulating synaptic decay (33). More information on DCM is in the supplementary methods. Leave-one-out cross-validation assessed whether connectivity patterns could classify the left-out participant as having a depression history or reaching the BDI-II and HADS-D clinical cut-off.

## 5. Results

### 5.1. General characteristics

HDGECs did not differ in sex, age, ethnicity, handedness, or HD-related variables. HDGECs with a history of depression had significantly higher history of suicidal ideation, use of mood medication and HADS-D scores. Participant characteristics are summarized in Table 1.

**Table 1.**
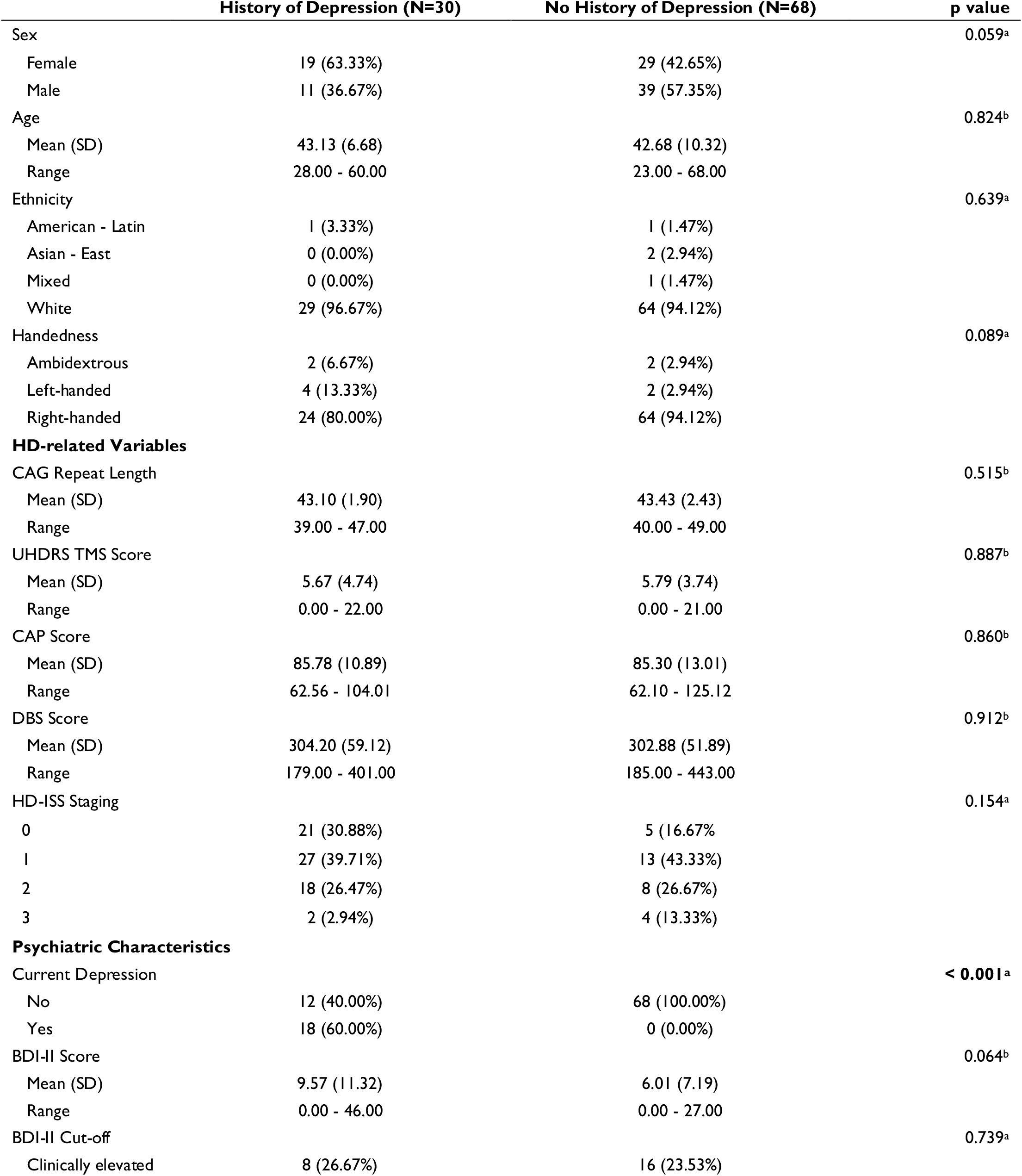
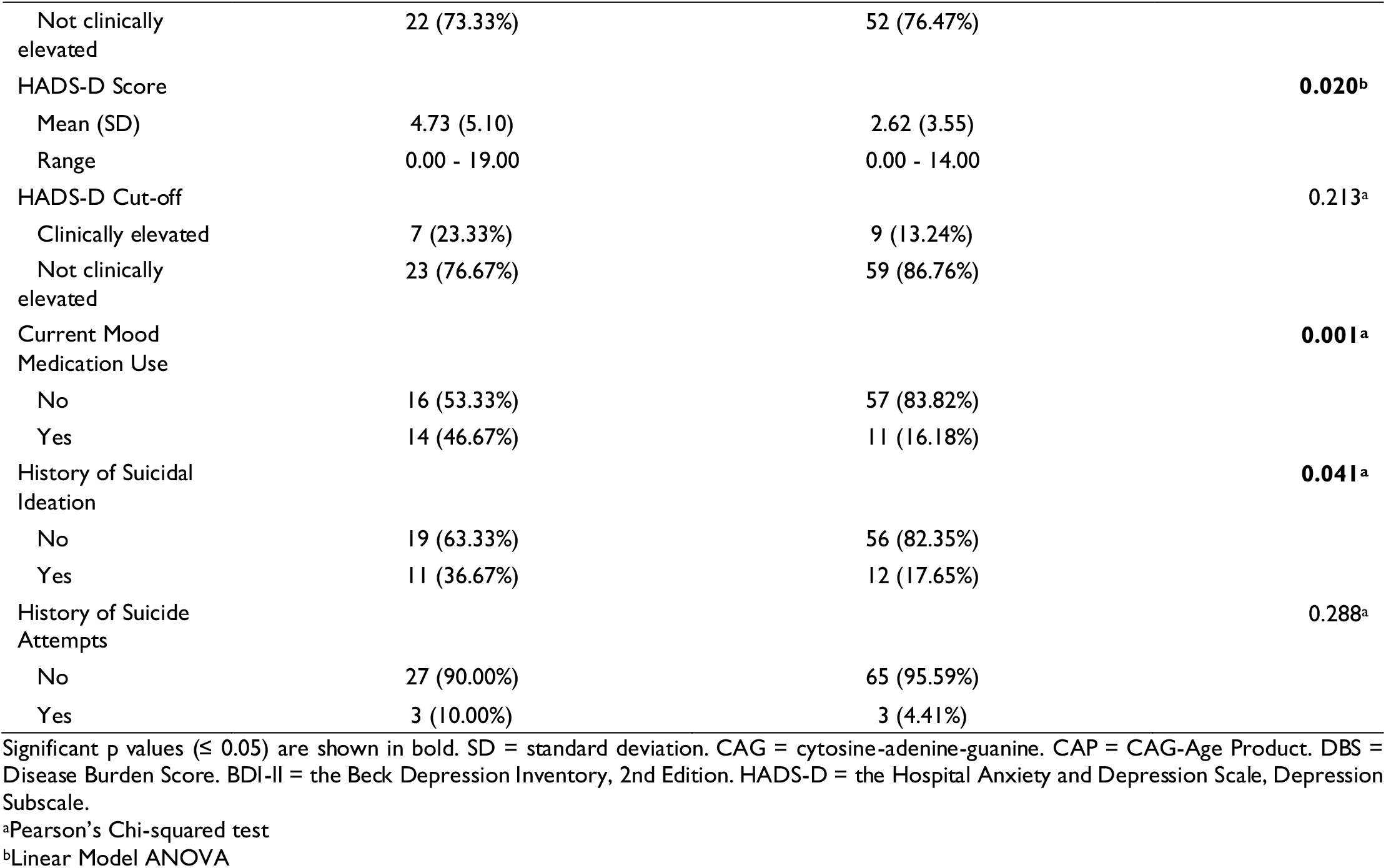
Demographic, Clinical, and Psychiatric Characteristics of Participant Groups.

### 5.2. Accuracy of model estimation

The estimation of DCM models for individual participants was excellent, with average variance-explained of 89.82% (*SD* = 3.89; *range* = 75.15-96.44; eFig. 9).

### 5.3. Effective Connectivity Differences Between HDGECs with a History of Depression and Those Without

For HDGECs with a history of depression compared to those without (Fig. 2), there was greater involvement of posterior DMN. This included disinhibitory PCC efferent to both hippocampi, right hippocampal increased inhibition to PCC, and left hippocampal increased inhibition to MPFC. Within striatum, inhibitory self-connectivity of bilateral caudate increased. Additionally, there was increased inhibition of left caudate on left putamen.

**Fig. 2:**
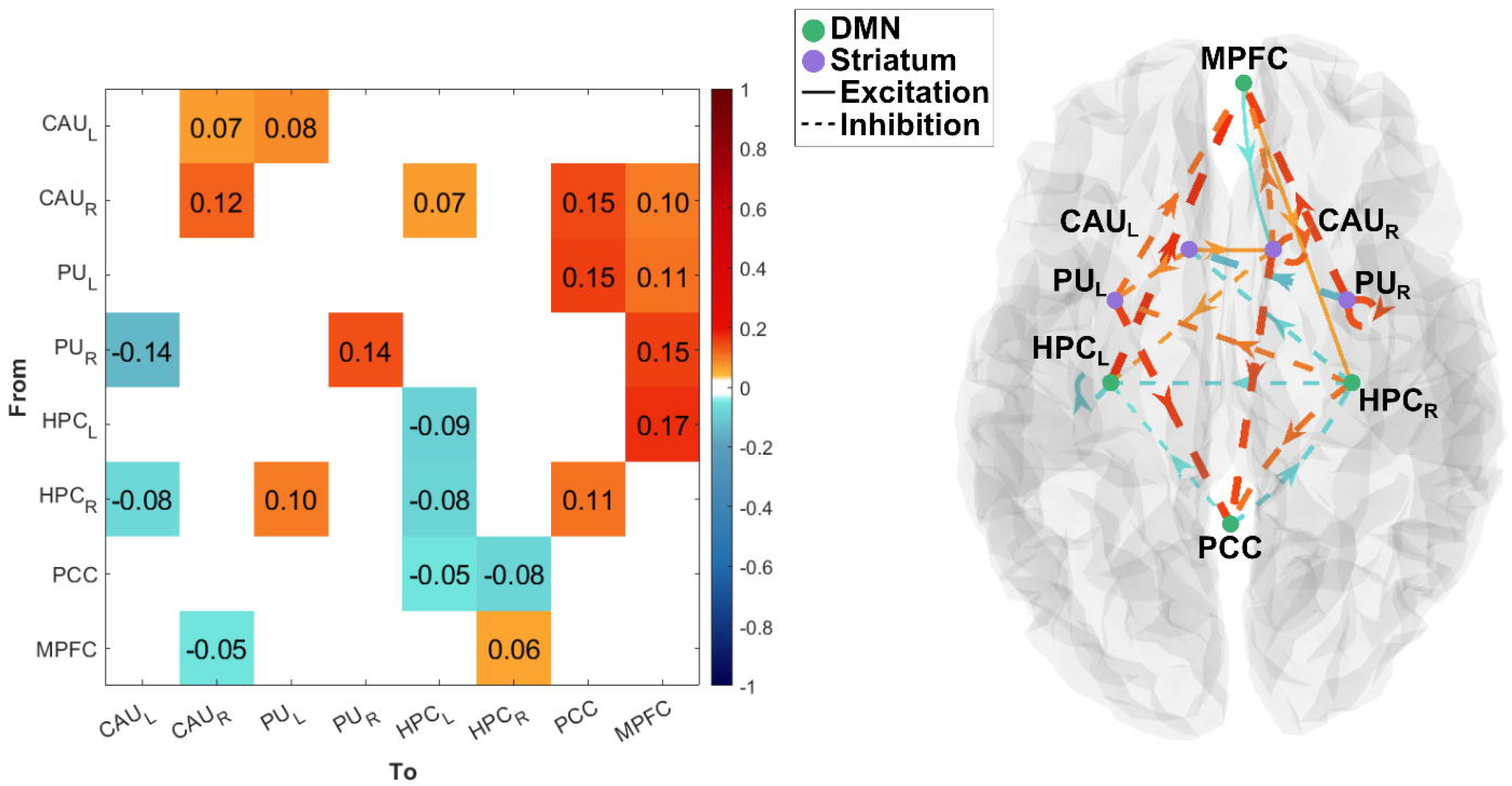
Difference in Mean Effective Connectivity Between HDGECs with a History of Depression Versus Without. **(L)** Connectivity matrix with increased connectivity in yellow-red and decreased connectivity in aqua-navy. **(R)** Effective connections that reached significance are shown in the dorsal plane. Nodes in purple represent the striatum, while those in green represent the default mode network. Weighted edges and arrows show the strength and directed influence of one region on another, including self-connections. Solid lines represent excitatory connections while dashes represent inhibitory connections. MPFC = Medial Prefrontal Cortex; PCC = Posterior Cingulate Cortex; HPC_L_ = Left Hippocampus; HPC_R_ = Right Hippocampus; CAU_L_ = Left Caudate; CAU_R_ = Left Caudate; PU_L_ = Left Putamen; PU_R_ = Right Putamen.

Between-network interactions revealed the striatum had increased inhibitory influence on MPFC and PCC for HDGECs with a history of depression. The MPFC had decreased excitatory influence on right caudate, right hippocampus showed disinhibition to left caudate and increased inhibition to left putamen (see eTable 1 for credible intervals). We ran sub-analyses using a covariate for each medication class: antipsychotics, benzodiazepines, SSRIs and non-SSRI, and another only including participants not using any mood medications. In both sub-analyses, we found concordant patterns of connectivity changes (see supplementary results).

Several connections accurately classified HDGECs depression history status, including right putamen to left caudate (*p* =.008) and MPFC (*p* =.032), PCC to right hippocampus (*p* =.02), as well as from PCC to bilateral hippocampi (*p* =.023) and within-DMN connections (*p* =.046). See eTable 4 and 5.

#### 5.3.1. Brain-behavior Associations

We examined effective connectivity changes associated with clinically elevated HADS-D and BDI-II scores, for HDGECs with and without a history of depression (Fig. 3).

**Fig. 3:**
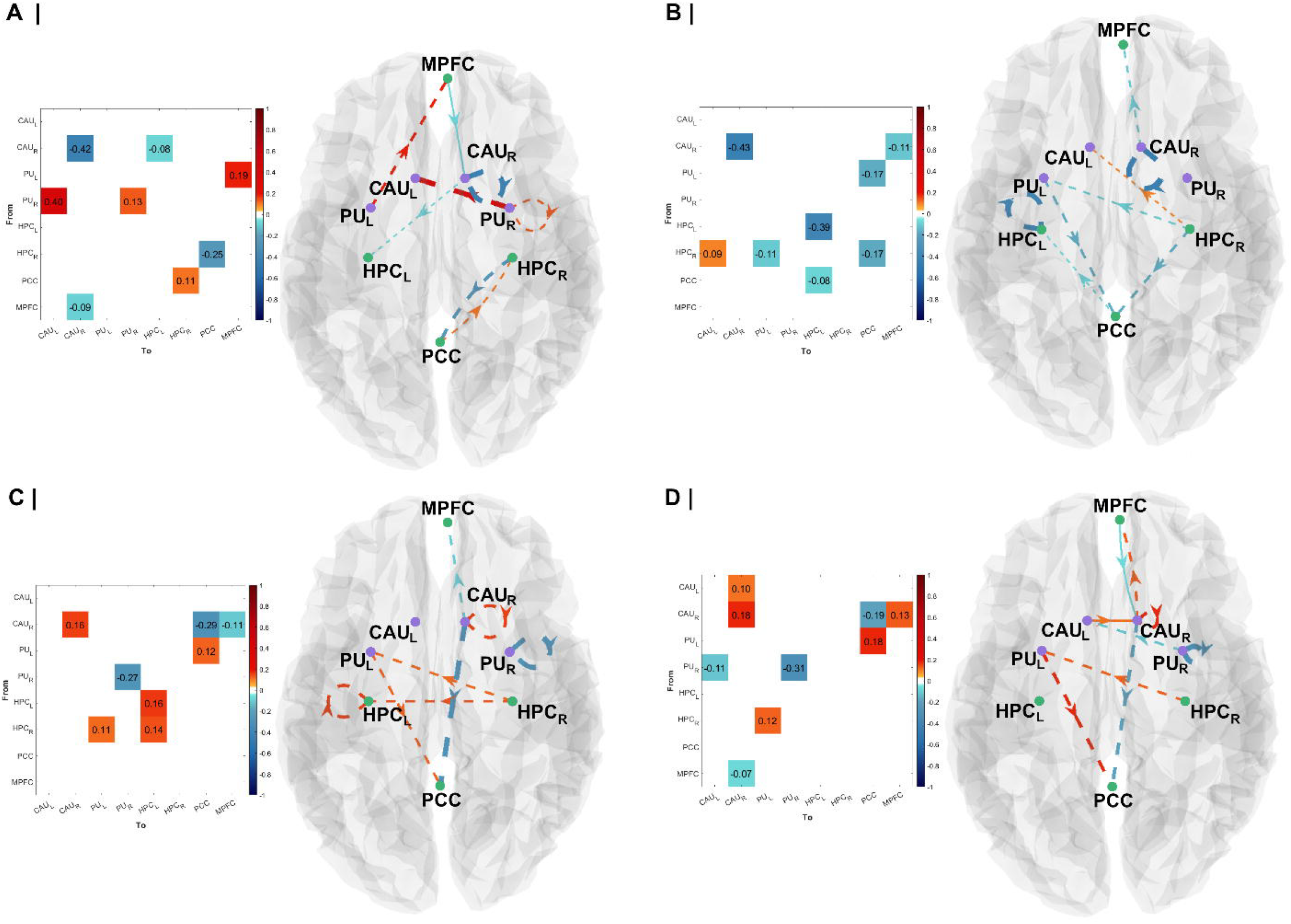
Effective Connectivity of DMN And Striatum in Association with Meeting Clinical Cut-Off for HADS-D And BDI-II. **(A)** Regional connectivity changes for HDGECs with depression history associated with meeting clinical cut-off for BDI-II. **(B)** Regional connectivity changes for HDGECs with depression history associated with meeting clinical cut-off for HADS-D. **(C)** Regional connectivity changes for HDGECs with no depression history associated with meeting clinical cut-off for BDI-II. **(D)** Regional connectivity changes for HDGECs with no depression history associated with meeting clinical cut-off for HADS-D. Weighted edges and arrows show the strength and directed influence of one region on another, including self-connections. Solid lines represent excitatory connections, dashed inhibitory, from the group difference DCM analysis. MPFC = Medial Prefrontal Cortex; PCC = Posterior Cingulate Cortex; HPC_L_ = Left Hippocampus; HPC_R_ = Right Hippocampus; CAU_L_ = Left Caudate; CAU_R_ = Left Caudate; PU_L_ = Left Putamen; PU_R_ = Right Putamen.

For HDGECs with a history of depression, clinical measures revealed alterations in DMN connectivity: meeting the clinical cut-off for BDI-II (Fig. 3A) and HADS-D (Fig. 3B) was negatively associated with right hippocampus to PCC connectivity. Within the striatum, there negative association with right caudate self-connectivity. Between-network interactions showed measure-specific patterns: meeting the HADS-D clinical cut-off was negatively associated with right hippocampus to left putamen connectivity, while meeting the BDI-II clinical cut-off was negatively associated with left putamen to MPFC connectivity.

For HDGECs without a history of depression, meeting the BDI-II clinical cut-off (Fig. 3C) was positively associated with right to left hippocampus and left hippocampal self-connectivity, while meeting the HADS-D clinical cut-off (Fig. 3D) showed no associations with DMN connectivity. Within the striatum, meeting the clinical cut-off for both measures was positively associated with right caudate self-connectivity and negatively associated with right putamen self-connectivity. Between-network interactions showed consistency: meeting the clinical cut-off was negatively associated with right caudate to PCC connectivity and positively associated with left putamen to PCC and right hippocampus to left putamen connectivity.

Effective connectivity from right putamen to left caudate classified HDGECs who had elevated depressive symptoms on BDI-II, (*p* =.006; eTable 5). No connections predicted HADS-D scores.

### 5.4. Current and Remitted Depression Effective Connectivity Differences

#### 5.4.1. Current Versus Never Depressed

When comparing HDGECs with currently diagnosed depression (*n* = 18) to HDGECs with no history of depression, (*n* = 68; Fig. 4A), many connections remained significant with stronger increases and decreases in connectivity. Key differences were the loss of influence of MPFC and additional influence of the striatum, including left putamen efferent self-connectivity and to right hippocampus, right putamen to PCC connectivity, and left caudate to right putamen.

**Fig. 4:**
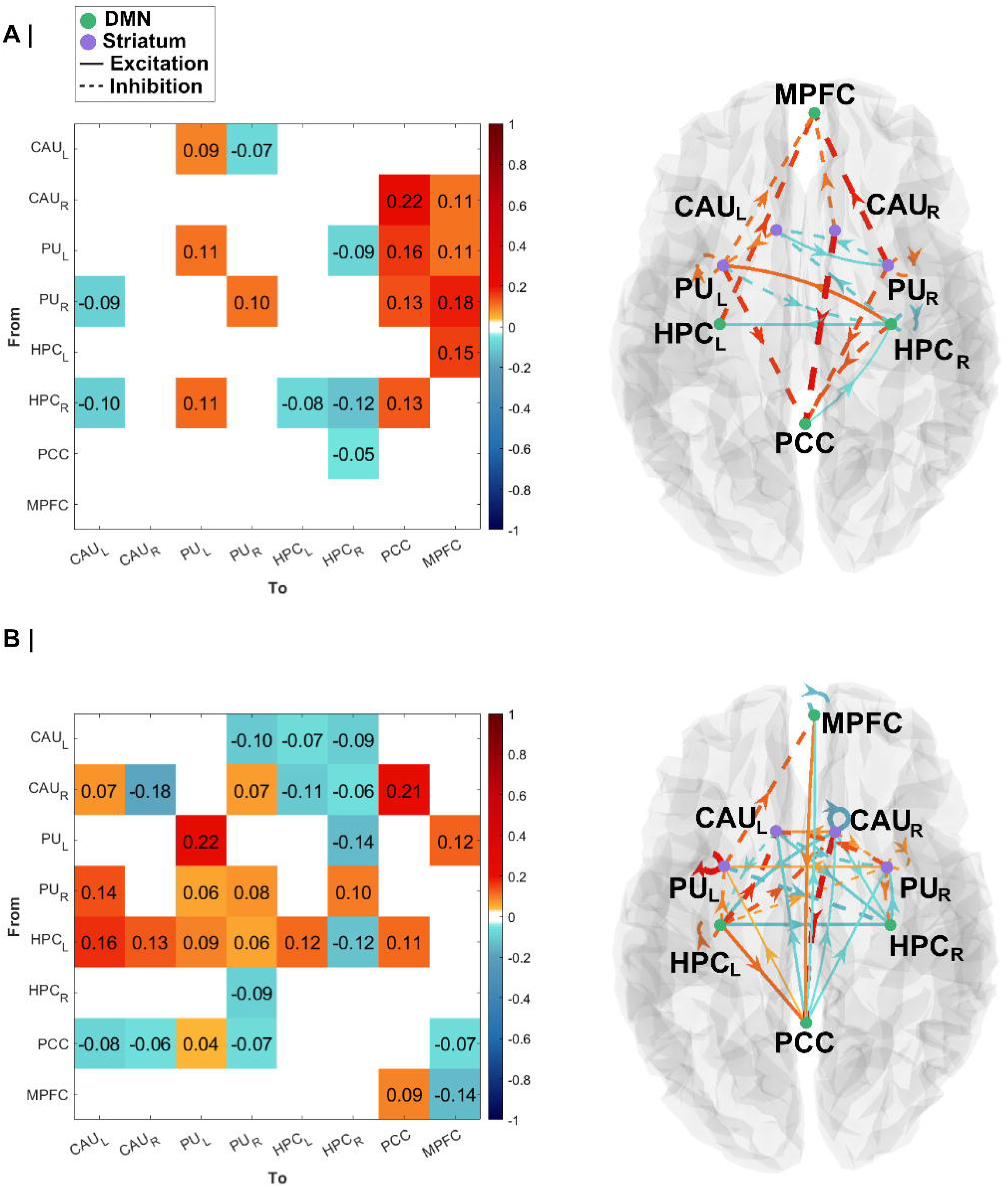
Mean Effective Connectivity Between HDGECs with Current Depression versus no Depression History and Current Depression versus Remitted. **(A)** Left panel shows connectivity matrix for current depression (with no depression as the reference), with increased connectivity in yellow-red and decreased connectivity in aqua-navy and right panel shows effective connections that reached significance are shown in the dorsal plane. **(B)** Left panel shows the connectivity matrix for matrix for current depression (with remitted depression as the reference), with increased connectivity in yellow-red and decreased connectivity in aqua-navy and right panel shows effective connections that reached significance are shown in the dorsal plane. Nodes in purple represent the striatum, green represents the default mode network. Weighted edges and arrows show the strength and directed influence of one region on another, including self-connections. Solid lines represent excitatory connections while dashes represent inhibitory connections. MPFC = Medial Prefrontal Cortex; PCC = Posterior Cingulate Cortex; HPC_L_ = Left Hippocampus; HPC_R_ = Right Hippocampus; CAU_L_ = Left Caudate; CAU_R_ = Left Caudate; PU_L_ = Left Putamen; PU_R_ = Right Putamen

#### 5.4.2. Current Versus Remitted Depression

There were unique connectivity aberrations for currently depressed HDGECs (*n* = 18) compared to HDGECs with remitted depression (*n* = 12; Fig. 4B). The MPFC had increased connectivity to PCC while PCC had decreased connectivity to MPFC, and we saw MPFC self-connectivity changes. PCC had decreased influence across striatal regions, except left putamen. Left hippocampus had increased connectivity with both the striatum and DMN. Additionally, caudate connectivity was more influential, particularly right caudate.

## 6. Discussion

Here, we show novel patterns of network dysfunction associated with depression for premanifest HDGECs. As hypothesized, HDGECs with a history of depression had increased excitatory connectivity from medial prefrontal to hippocampus and decreased excitatory connectivity from MPFC to striatum. Contrary to predictions, MPFC to PCC connectivity did not increase; instead, we show increased inhibitory striatal-to-DMN connectivity. Within the DMN, inhibitory interactions between hippocampal regions and PCC were altered, suggesting complex reorganization. For HDGECs with a depression history, clinically significant depressive symptoms were negatively associated with caudate self-connectivity, which was negative for those without. Additionally, having clinically elevated depression was negatively associated with right hippocampus to PCC connectivity for HDGECs with depression history, or right caudate to PCC for those without a history of depression. When comparing current versus remitted depression, we observed the expected increased influence of MPFC on PCC and decreased connectivity from PCC to MPFC.

The present findings establish distinct patterns of striatal-DMN interactions associated with depressive symptoms for HDGECs. Notably, while striatal dysfunction is a hallmark of HD in general (9), we found unique patterns of striatal connectivity between groups, suggesting depression-specific dysfunction. Our findings of left putamen involvement is consistent with structural neuroimaging evidence of leftward putamen grey matter loss associated with emotion processing (62). The current study extends these structural asymmetry findings by revealing striatal regions have increased inhibitory influence on DMN in association with depression history for HDGECs. This suggests left putamen’s structural vulnerability is accompanied by functional changes in broader striatal connectivity in the context of depression in HDGECs. Previous investigations in the same cohort (using a whole-brain approach) showed increased functional connectivity between cingulate, medial orbitofrontal, and parahippocampal regions, but not in striatal regions (27). Our approach reveals that the DMN shows complex patterns of increasing and decreasing communication with the striatum as well as within-striatum increased connectivity. Additionally, right caudate self-connectivity associated differently between groups when looking at depression symptom severity. This shows how DCM detects depression-related directional striatal communication changes missed by whole-brain approaches. When comparing current versus never-depressed HDGECs, consistent differences in connection strength and direction further support a depression-specific nature. These patterns, particularly putamen-DMN connectivity, could potentially serve as markers of depression for HDGECs.

These findings link hippocampal dysconnectivity and depression for HDGECs. In keeping with depression in non-neurological populations, there was a propensity for right hippocampal involvement (63,64), however, our findings reveal differences compared to major depression. While major depression is associated with increased right hippocampal functional connectivity (63), our findings demonstrate increasing and decreasing inhibitory right hippocampal connections, suggesting depression for HDGECs involves more complex interactions. Moreover, decreased inhibitory PCC-hippocampal connectivity patterns are in keeping with findings of the PCC as a disease epicenter for HD (15), and our findings suggest that posterior DMN regions are drivers of depression for HDGECs. The prominent hippocampal connectivity changes may reflect multiple mechanisms. Hippocampal dysfunction is shown in non-neurological depression (63,64) and existing HDGEC research has identified broader parahippocampal connectivity changes (27), therefore, alterations may represent shared depression mechanisms between HD and major depression. It is possible that hippocampal dysconnectivity represents functional reorganization. This interpretation aligns with broader evidence for pathological hyperconnectivity and compensation in HDGECs, with increased functional connectivity between preserved regions (including hippocampus, PCC and frontal regions)(15,16,37). Notably, patterns predominantly involve the right hemisphere (16), consistent with our finding of prominent right hippocampal involvement, providing supplementary evidence to support a role for the hippocampus in depression for HDGECs.

A striking finding was the limited influence of MPFC. Increased inhibitory influence of right caudate on MPFC extends existing literature of hyperconnectivity of MPFC and caudate in major depression (22,23). Aberrant connectivity between these regions is driven by striatal efferents, which functional connectivity techniques did not elucidate. When examining current versus remitted depression, hypothesized increased MPFC to PCC connectivity was observed, suggesting state-dependent MPFC dysfunction. In major depression, a treatment-resistant DMN subnetwork includes resolved posterior functional dysconnectivity alongside continued aberrant MPFC connectivity with treatment (24). By contrast, our findings showed differences between current and remitted states, particularly in MPFC connectivity and continued posterior DMN influence regardless of current or remitted depression. This suggests that depression in HDGECs is driven by posterior DMN changes and may be distinguishable from major depression. It is possible that the limited influence of MPFC could be attributed to the choice of prefrontal region. The division of the MPFC into ventral and dorsal regions is common, with no definition or number of subdivisions (65,66), and anterior DMN has included orbitofrontal, ventromedial, and dorsomedial cortices (67,68). It is possible that our study did not fully capture the anterior DMN hub, however, supplementary analyses found that excitatory influence of MPFC on PCC and disinhibition of MPFC cancelled out at the group level. This suggests that these connections do not distinguish HDGECs with depression from those without.

We found unique connectivity changes across BDI-II and HADS-D and it’s possible we did not capture the most clinically relevant depression symptoms. Most major depression criteria include somatic items (e.g., sleep disturbances, psychomotor changes) that lack diagnostic specificity in HDGECs, as they are also HD symptoms unrelated to depression (10). For example, the HADS-D has better sensitivity and specificity in HD compared to the BDI-II because it has only one somatic item (52). Despite this, endorsed depressive symptoms by HDGECs are largely consistent with standard diagnostic criteria (10). Therefore, both measures and clinical cut-offs were used as the most meaningful measure of depressive symptoms. Additionally, the effect of mood medications on the BOLD signal presented another challenge. Cerebrovascular changes are reported with acute mood medication use, including cerebral blood flow changes with antipsychotics (69,70), and SSRIs (71,72). This is further complicated by changes in cerebral blood flow as a function of major depression itself (73). Having the covariate and using DCM (which includes a neurovascular model) helped address this limitation. Sub-analyses showing similar findings regardless of how we controlled for mood medication suggest our parsimonious approach captured neuronal differences and our findings are compared to research with similar methodologies (16,27,37–39,62).

These findings motivate future directions. The temporal pattern of depression in HD remains contentious. Studies suggest depression is unrelated to CAG repeat length (8,74,75) and disease progression (2,74,76), however, it is linked to cognitive and motor changes (77) and remains elevated in manifest HD (78,79). This suggests a mechanistic relationship between HD progression and depression. Broadly, there is transition from functional hyperconnectivity for HDGECs to hypoconnectivity as disease burden increases (15,16). Therefore, there remains an unresolved question about effective connectivity changes associated with depression in HDGECs over disease progression.

Overall, our findings suggest DMN and striatal dysconnection as a neural basis for depression in HDGECs. Aberrant effective connections were associated with diagnosed depression history, which was differentially associated with coupling changes in depressive symptoms. Defining functional brain networks of neuropsychiatric features, including depression, plays an important role in understanding the pathophysiology of HD.

## Data Availability

No new data were collected for this article. The data that support the findings of this study are available on the Enroll-HD platform.

https://www.enroll-hd.org/for-researchers/access-data-biosamples/

## 7. Supplementary material

Supplementary material is available online.

## Notes

AR was funded by the Australian National Health and Medical Research Council (Investigator Grant 1194910) for this work. TB was funded by Research Training Program (RTP) scholarship for this work. There were no specific sources of funding for this work. The authors declare that there are no conflicts of interest relevant to this work.

### Competing Interest Statement

The authors have declared no competing interest.

### Funding Statement

AR and LN are funded by the Australian Research Council (DP200100757) and AR by Australian National Health and Medical Research Council Investigator Grant (Ref: 1194910). AR is affiliated with The Wellcome Centre for Human Neuroimaging supported by core funding from Wellcome [203147/Z/16/Z]. AR is a CIFAR Azrieli Global Scholar in the Brain, Mind, and Consciousness Program.

### Author Declarations

This study used data from Track-On HD, which was approved by local ethics committees at all four participating sites: Leiden (Netherlands), London (United Kingdom), Paris (France), and Vancouver (Canada), and has been reported in Kloppel et al. (2015) and Tabrizi et al. (2009).

### Summary of Updates

Sections on medication use and depression history updated, additional medication use subset analyses completed. Figures 1-4 all updated, supplemental files updated.

