## Supplementary Materials for "Depression in Premanifest Huntington’s Disease: Aberrant Effective Connectivity of Striatum and Default Mode Network"

Author affiliations:

### Methods

#### Imaging Data Acquisition

3T MRI data was acquired on two different scanner systems, Philips Achieva at Leiden and Vancouver and Siemens TIM Trio at London and Paris, described in detail elsewhere (1). For T1-weighted image acquisition, a 3D MPRAGE sequence was used with the following parameters: TR 2200ms (Siemens)/ 7.7ms (Philips), TE 2.2ms (S)/3.5ms (P), FOV 280 mm (S)/240 mm (P), flip angle 10°(S)/8°(P), 208(S)/164(P) sagittal slices (slice thickness: 1.1 mm, gap: no gap, matrix size 256 × 256 (S)/224 × 224 (P)) and bandwidth of 240 Hz (S)/241 Hz (P) per participant (2). For whole-brain volume acquisition, a T2-weighted echo planar imaging sequence was used with the following parameters: TR 3000 ms, TE 30 ms, FOV 212 mm, flip angle 80°, 48 slices in ascending order (slice thickness: 2.8 mm, gap: 1.5 mm, in plane resolution 3 × 3 × 3 mm) and bandwidth of 1906 Hz per participant (1).

#### Pre-processing

Firstly data was reorganised to BIDS specification to standardise the data structure, ensure dataset integrity, and define the readable metadata (3). Preprocessing was performed for all participants using *fMRIPrep* 21.0.2 (Esteban, Markiewicz, et al. (2018); Esteban, Blair, et al. (2018); RRID:SCR_016216) (4,5)—described in detail below—and MRIQC v22.0.6 (6), which both used FreeSurfer v-6.0.1 (7). Spatial smoothing by a 6 mm full-width half-maximum Gaussian kernel was subsequently undertaken using SPM12 (7771). MRIQC is a quality control tool for anatomical and fMRI data that extracts no-reference image quality metrics for scan quality assessments (6). It is used to compliment fMRIPrep and provided group level quality control assessments. Pre-processed derivatives from fMRIPrep provided the input for the subsequent DCM analyses.

#### fMRIPrep

Results included in this manuscript come from preprocessing performed using *fMRIPrep* 21.0.2 (8,9)(RRID:SCR_016216), which is based on *Nipype* 1.6.1 (10,11) (RRID:SCR_002502).

##### Anatomical data preprocessing

A total of 3 T1-weighted (T1w) images were found within the input BIDS dataset. All of them were corrected for intensity non-uniformity (INU) with N4BiasFieldCorrection (12), distributed with ANTs 2.3.3 (13)(RRID:SCR_004757). The T1w-reference was then skull-stripped with a *Nipype* implementation of the antsBrainExtraction.sh workflow (from ANTs), using OASIS30ANTs as target template. Brain tissue segmentation of cerebrospinal fluid (CSF), white-matter (WM) and gray-matter (GM) was performed on the brain-extracted T1w using fast (FSL 6.0.5.1:57b01774, RRID:SCR_002823) (14). A T1w-reference map was computed after registration of 3 T1w images (after INU-correction) using mri_R_obust_template (FreeSurfer 6.0.1)(15). Brain surfaces were reconstructed using recon-all (FreeSurfer 6.0.1, RRID:SCR_001847)(16), and the brain mask estimated previously was refined with a custom variation of the method to reconcile ANTs-derived and FreeSurfer-derived segmentations of the cortical gray-matter of Mindboggle (RRID:SCR_002438)(17). Volume-based spatial normalization to one standard space (MNI152NLin2009cAsym) was performed through nonlinear registration with antsRegistration (ANTs 2.3.3), using brain-extracted versions of both T1w reference and the T1w template. The following template was selected for spatial normalization: *ICBM 152 Nonlinear Asymmetrical template version 2009c* [RRID:SCR_008796; TemplateFlow ID: MNI152NLin2009cAsym](18).

##### Functional data preprocessing

For each of the 3 BOLD runs found per subject (across all tasks and sessions), the following preprocessing was performed. First, a reference volume and its skull-stripped version were generated using a custom methodology of *fMRIPrep*. Head-motion parameters with respect to the BOLD reference (transformation matrices, and six corresponding rotation and translation parameters) are estimated before any spatiotemporal filtering using mcflirt (FSL 6.0.5.1:57b01774)(19). BOLD runs were slice-time corrected to 1.48s (0.5 of slice acquisition range 0s-2.95s) using 3dTshift from AFNI (20)(RRID:SCR_005927). The BOLD time-series (including slice-timing correction when applied) were resampled onto their original, native space by applying the transforms to correct for head-motion. These resampled BOLD time-series will be referred to as *preprocessed BOLD in original space*, or just *preprocessed BOLD*. The BOLD reference was then co-registered to the T1w reference using bbregister (FreeSurfer) which implements boundary-based registration (21). Co-registration was configured with six degrees of freedom. Several confounding time-series were calculated based on the *preprocessed BOLD*: framewise displacement (FD), DVARS and three region-wise global signals. FD was computed using two formulations following Power (absolute sum of relative motions (22), and Jenkinson (relative root mean square displacement between affines (19)). FD and DVARS are calculated for each functional run, both using their implementations in *Nipype* (following the definitions by Power et al. 2014). The three global signals are extracted within the CSF, the WM, and the whole-brain masks. Additionally, a set of physiological regressors were extracted to allow for component-based noise correction (*CompCor*, (23)). Principal components are estimated after high-pass filtering the *preprocessed BOLD* time-series (using a discrete cosine filter with 128s cut-off) for the two *CompCor* variants: temporal (tCompCor) and anatomical (aCompCor). tCompCor components are then calculated from the top 2% variable voxels within the brain mask. For aCompCor, three probabilistic masks (CSF, WM and combined CSF+WM) are generated in anatomical space. The implementation differs from that of Behzadi et al. in that instead of eroding the masks by 2 pixels on BOLD space, the aCompCor masks are subtracted a mask of pixels that likely contain a volume fraction of GM. This mask is obtained by dilating a GM mask extracted from the FreeSurfer’s *aseg* segmentation, and it ensures components are not extracted from voxels containing a minimal fraction of GM. Finally, these masks are resampled into BOLD space and binarized by thresholding at 0.99 (as in the original implementation). Components are also calculated separately within the WM and CSF masks. For each CompCor decomposition, the *k* components with the largest singular values are retained, such that the retained components’ time series are sufficient to explain 50 percent of variance across the nuisance mask (CSF, WM, combined, or temporal). The remaining components are dropped from consideration. The head-motion estimates calculated in the correction step were also placed within the corresponding confounds file. The confound time series derived from head motion estimates and global signals were expanded with the inclusion of temporal derivatives and quadratic terms for each (24). Frames that exceeded a threshold of 0.5 mm FD or 1.5 standardised DVARS were annotated as motion outliers. The BOLD time-series were resampled into standard space, generating a *preprocessed BOLD run in MNI152NLin2009cAsym space*. First, a reference volume and its skull-stripped version were generated using a custom methodology of *fMRIPrep*. All resamplings can be performed with *a single interpolation step* by composing all the pertinent transformations (i.e. head-motion transform matrices, susceptibility distortion correction when available, and co-registrations to anatomical and output spaces). Gridded (volumetric) resamplings were performed using antsApplyTransforms (ANTs), configured with Lanczos interpolation to minimize the smoothing effects of other kernels (Lanczos 1964). Non-gridded (surface) resamplings were performed using mri_vol2surf (FreeSurfer).

Many internal operations of *fMRIPrep* use *Nilearn* 0.8.1 (25)(RRID:SCR_001362), mostly within the functional processing workflow. For more details of the pipeline, see [the section corresponding to workflows in *fMRIPrep*’s documentation](https://fmriprep.readthedocs.io/en/latest/workflows.html).

##### Copyright Waiver

The above boilerplate text was automatically generated by fMRIPrep with the express intention that users should copy and paste this text into their manuscripts *unchanged*. It is released under the [CC0](https://creativecommons.org/publicdomain/zero/1.0/) license.

### Statistical Analyses

#### Spectral Dynamic Causal Modelling

Dynamic causal modelling (DCM), a popular method to infer effective connectivity, is a series of modelling techniques that are biophysically informed effective connectivity analyses of distributed neural networks (26) .DCM broadly refers to the Bayesian modelling procedure, which allows the testing of each potential model and its corresponding hypothesis regarding the construction of the network (26,27). In this way, it is a hypothesis-driven approach to understanding the neural networks underpinning observed brain activation (27). Model formulation evaluates how data are caused in the network, and DCM uses parameterised connections to understand the dynamics of intrinsic hidden states in the sources of fMRI data (26). Model inversion allows neuronal states to influence other neuronal states (26). DCM facilitates a mechanistic approach to understanding distributed neural processing and disturbances in connectivity changes in a pre-specified network of brain regions (26,27). There are two key forms of DCM used for rs-fMRI: stochastic DCM and spectral DCM (spDCM).

To model resting-state activity, which occurs in the absence of external stimuli, we incorporate a stochastic component to account for neural fluctuations within the model. Mathematically, we express the formulation of the stochastic generative model through two equations. The first equation represents the neuronal state dynamics as follows:

$$x˙\left( t \right)=f\left( x\left( t \right),u\left( t \right),\theta\right)+v\left( t \right) (1)$$

The second equation represents the observation process, which is a static nonlinear mapping from the hidden physiological states in equation (1) to the observed BOLD activity:

$$y\left( t \right)=h\left( x\left( t \right),\phi\right)+e\left( t \right) (2)$$

Here, *x*˙(*t*) represents the rate of change of the neuronal states *x*(*t*), where *θ* denotes the unknown parameters, specifically the effective connectivity. The terms $v\left( t \right)$ and $e\left( t \right)$ correspond to stochastic processes referred to as state noise and measurement (or observation) noise, respectively. These processes model the random neuronal fluctuations responsible for driving the resting-state activity.

In equation (2), $\phi$ represents the unknown parameters associated with the hemodynamic observation function. Additionally, $u\left( t \right)$ signifies any exogenous (or experimental) inputs that influence the hidden states. It's important to note that such inputs are typically absent in resting-state designs.

Criticisms of using stochastic DCM for fMRI data include its unstable model inversion and high computational cost by accounting for neuronal variation in the time domain (28,29). The use of spDCM ensures stability in estimations and computational efficiency (28,29). Research has demonstrated that spDCM provides more accurate estimates that are sensitive to group differences, when compared to stochastic DCM (29).

spDCM introduces a constrained inversion approach to the stochastic model by parameterizing the neuronal fluctuations $v\left( t \right)$. This methodology simplifies the generative model by substituting the original time series with their second-order statistics, specifically the cross spectra. This implies that instead of estimating time-varying hidden states, we are now focused on estimating their time-invariant covariance. To achieve this, we need to estimate the covariance of the random fluctuations. In this context, a scale-free (power-law) form for the state noise (or observation noise) is employed:

$$g_{v}\left( \omega,\theta\right)=\alpha_{v}\omega{}^{-\beta v}$$

$$g_{e}\left( \omega,\theta\right)=\alpha_{e}\omega^{-\beta e} (3)$$

In these equations, the parameters $\{\alpha,\beta\}\subset\theta$ govern the amplitudes and exponents of the spectral density of the neural fluctuations. Notably, the parameterization of endogenous fluctuations results in deterministic states, simplifying the inversion scheme significantly. Consequently, the estimation process now focuses solely on the model's parameters and hyperparameters.

#### Parametric Empirical Bayes

Empirical Bayes refers to the Bayesian inversion or fitting of hierarchical models. In hierarchical models, constraints on the posterior density over model parameters at any given level are determined by the level above, and these constraints are referred to as empirical priors as they are informed by empirical data. The hierarchical parametric empirical Bayes (PEB) model for DCM parameters elucidates how individual (within-subject) connections are derived from group membership. Parametric random effects modelling utilises the complete posterior density over the parameters from each participant's DCM, encompassing both the expected strength of each connection and the associated uncertainty (i.e., posterior covariance), to inform the group-level result (i.e., group differences).

Mathematically, for DCM studies with *N* participants and *M* parameters per DCM, the responses of the *i*-th participant and the distribution of the parameters over participants can be modelled as:

$y_{i}=\Gamma_{i}^{\left( 1 \right)}\left( \theta^{\left( 1 \right)} \right)+ \varepsilon_{i}^{\left( 1 \right)}$

$\theta^{\left( 1 \right)}=\Gamma^{\left( 2 \right)}\left( \theta^{\left( 2 \right)} \right)+ \varepsilon^{\left( 2 \right)}$	(4)

$\theta^{\left( 2 \right)}=\eta+ \varepsilon^{\left( 3 \right)}$

Here, $y_{i}$ represents the BOLD time series from the *i*-th participant, and $\Gamma_{i}^{\left( 1 \right)}$ is a nonlinear mapping from the parameters of a model to the predicted response y, which in this study corresponds to the model in Eq. S1 above. $\varepsilon_{i}^{\left( 1 \right)}$represents independent and identically distributed observation noise (equivalent to $e\left( t \right)$ in Eq. S2).

$\Gamma^{\left( 2 \right)}\left( \theta^{\left( 2 \right)} \right)=(X\bigotimes W)\beta$	(5)

Here, $\beta\subset\theta$ represents group means or effects encoded by a design matrix comprising both between-subject $(X)$ and within-subject $\left( W \right)$components. The between-subject part encodes differences among subjects or covariates such as age, while the within-subject part specifies mixtures of parameters that exhibit random effects. It is assumed that the first column of the design matrix is a constant term that models group means, and subsequent columns encode group differences.

#### Self-Connectivity in Dynamic Causal Modelling

Within the DCM framework, self-connections are modelled as inhibitory to prevent potential runaway excitation; however, these self-connection parameters are logarithmically scaled (using the transformation log(−2*a*) in SPM). This logarithmic scaling is employed to enhance the numerical stability of the model fitting procedures and is technically motivated by using log-normal priors to enforce recurrent self-inhibition. Consequently, these self-connections can take both positive and negative values, with specific interpretations.

In SPM's reporting convention, a zero value (arbitrarily set) for a self-connection corresponds to −0.5 Hz, representing the default prior self-connectivity value. A positive self-connection signifies a relative increase in inhibition (faster decay rates, below 0.5 Hz), while a negative self-connection indicates a relative decrease in inhibition (slower decay rates in the −0.5 to 0 Hz range). These inhibitory self-connections regulate the gain or sensitivity to inputs from other regions. Decreased self-inhibition implies increased synaptic gain or sensitivity to inputs, whereas increased self-inhibition suggests a reduction in synaptic gain or sensitivity to inputs. Importantly, only self-connections are subjected to this logarithmic scaling transformation in DCM.

### Results

#### Participant Characteristics

##### Depression History

eTable 1 Depression History Classification Details

|  | History of depression (*n* = 30) |
| --- | --- |
| Remitted Depression | 12 (12.2%) |
| 1 remitted episode | 11 (91.67%) |
| 2 remitted episodes | 1 (8.33%) |
| Current Episode of Depression | 18 (18.4%) |
| First Episode of Depression | 17 (17.3%) |
| Recurrent Episode of Depression | 1 (1.0%) |
| Median Duration of Current Episode - years (IQR), range | 4 (2.3 - 6.7), 0.3 - 37.7 |
| ICD 10 Diagnostic Category |  |
| F32.9 (Depressive Episode, Unspecified) | 29 (96.7%) |
| F32.2 (Severe Depressive Episode without Psychotic Symptoms) | 1 (3.3%) |

**Note.** ICD = International Statistical Classification of Diseases and Related Health Problems 10th Revision

##### Mood Medication Use

eTable 2 Mood Medication Use Details

| Medication | N | *n* (history of depression) | *n* (no depression history) | Duration Mean (SD), range – years | Dose Mean (SD), range – mgs | Indication | Regime |
| --- | --- | --- | --- | --- | --- | --- | --- |
| **SSRI** |  |  |  |  |  |  |  |
| Citalopram | 11 | 5 | 6 | 2.11 (1.69), 0.40-5.73 | 20.9 (8.3), 10.0-40.0 | Anxiety (27%), Depression (73%) | once daily (100%) |
| Escitalopram | 2 | 1 | 1 | 2.44 (3.46), 0.00-4.89 | 10.0 (0.0), 10.0-10.0 | Depression (100%) | once daily (100%) |
| Fluoxetine | 1 | 1 | 0 | 3.87 (0.00), 3.87-3.87 | 20.0 (0.0), 20.0-20.0 | Depression (100%) | once daily (100%) |
| **Non-SSRI** |  |  |  |  |  |  |  |
| Amitriptyline | 1 | 1 | 0 | 13.78 (0.00), 13.78-13.78 | 50.0 (0.0), 50.0-50.0 | Post-traumatic stress (100%) | once daily (100%) |
| Bupropion | 1 | 0 | 1 | 0.66 (0.00), 0.66-0.66 | 150.0 (0.0), 150.0-150.0 | Anxiety (100%) | once daily (100%) |
| Mianserin | 1 | 0 | 1 | 0.41 (0.00), 0.41-0.41 | 30.0 (0.0), 30.0-30.0 | Depression (100%) | once daily (100%) |
| Mirtazapine | 1 | 1 | 0 | 1.04 (0.00), 1.04-1.04 | 15.0 (0.0), 15.0-15.0 | Depression (100%) | once daily (100%) |
| Venlafaxine | 2 | 2 | 0 | 0.74 (0.33), 0.51-0.98 | 93.8 (26.5), 75.0-112.5 | Depression (100%) | twice daily (50%), once daily (50%) |
| **Antipsychotic** |  |  |  |  |  |  |  |
| Aripiprazole | 1 | 1 | 0 | 0.31 (0.00), 0.31-0.31 | 15.0 (0.0), 15.0-15.0 | Depression (100%) | once daily (100%) |
| Olanzapine | 1 | 1 | 0 | 2.06 (0.00), 2.06-2.06 | 2.5 (0.0), 2.5-2.5 | Irritability (100%) | once daily (100%) |
| **Benzodiazepine** |  |  |  |  |  |  |  |
| Bromazepam | 1 | 0 | 1 | 3.24 (0.00), 3.24-3.24 | 1.5 (0.0), 1.5-1.5 | Insomnia (100%) | once daily (100%) |
| Oxazepam | 1 | 0 | 1 | 0.41 (0.00), 0.41-0.41 | 10.0 (0.0), 10.0-10.0 | Anxiety (100%) | twice daily (100%) |
| Temazepam | 1 | 1 | 0 | 14.83 (0.00), 14.83-14.83 | 10.0 (0.0), 10.0-10.0 | Sleep disorder (100%) | once daily (100%) |

**Note.** SSRI = Selective Serotonin Reuptake Inhibitors. All doses in mg, unless otherwise stated.

#### Assumptions Checks

For Pearson’s chi-squared tests, data used were nominal or ordinal, mutually exclusive, and groups were independent.

##### Linear Model ANOVA

###### Age

Residuals for were normally distributed (W = 0.992, *p* = 0.838), as supported by visual inspection of the Q-Q plot. The box plot showed no evidence of heteroscedasticity. See Supplementary Figure 1.

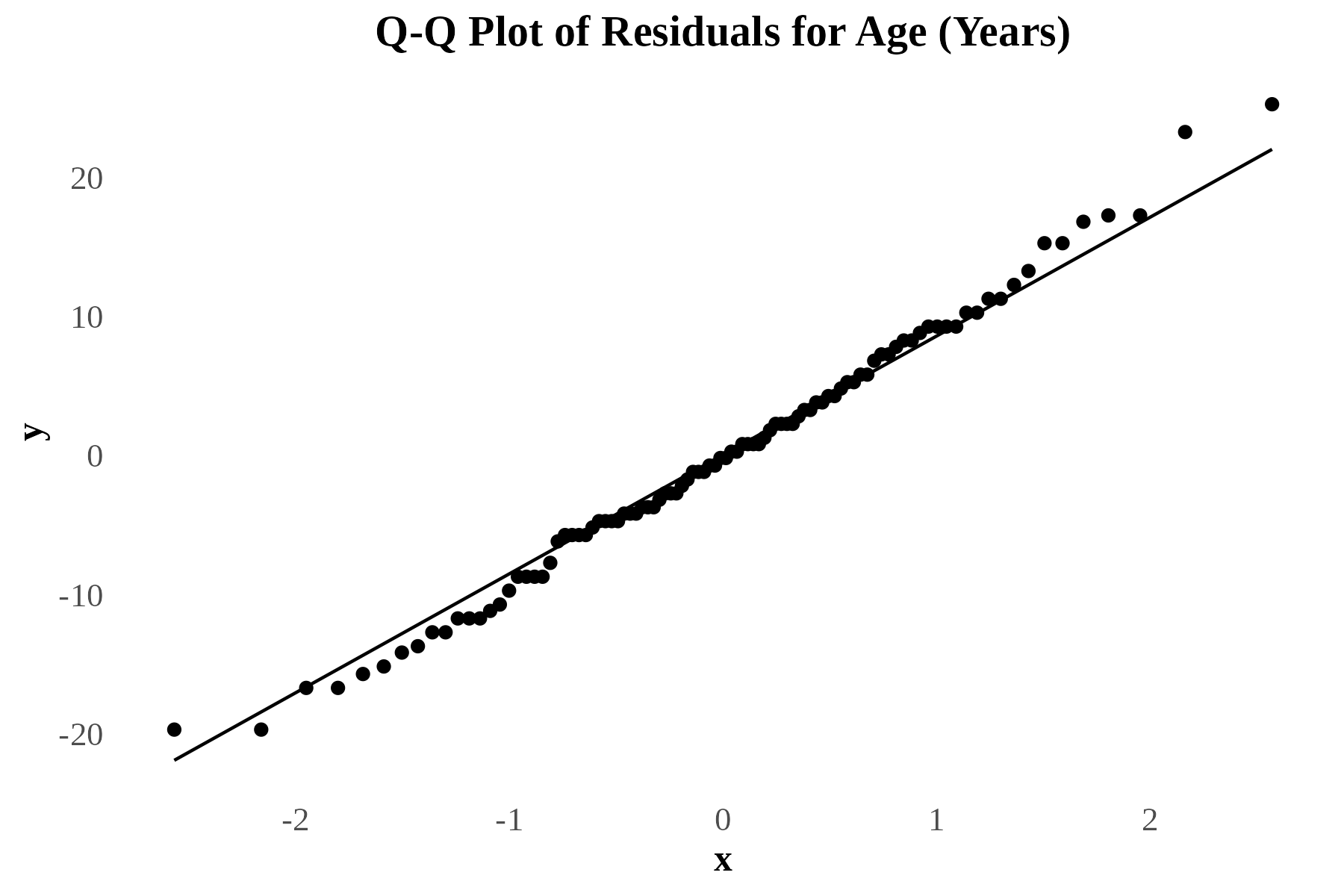

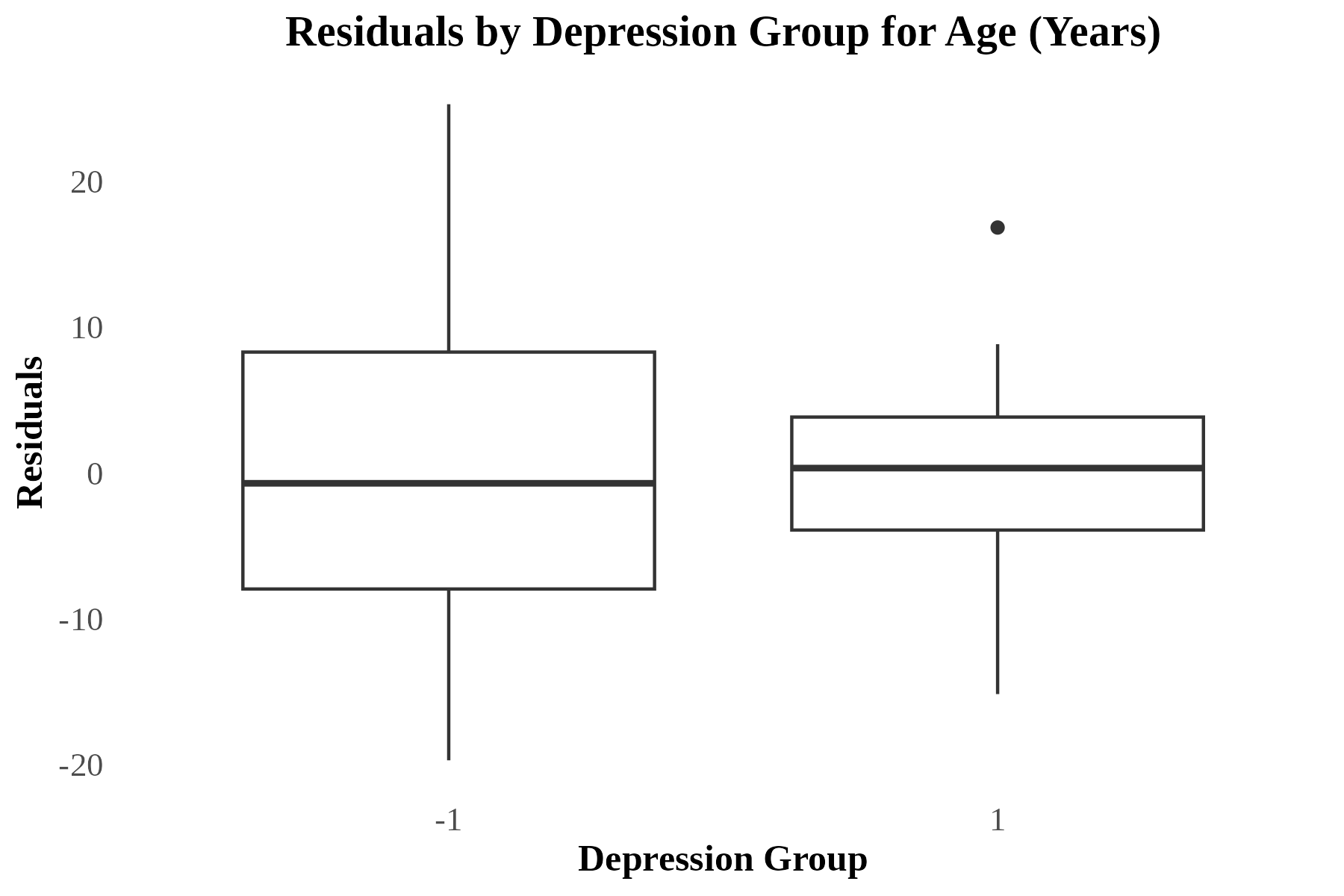

eFig. 1 Q-Q plot and boxplot of Residuals for Age. The left plot shows the results of the Q-Q plot of residuals for participant age. The right plot shows the results of the box plots of residuals for age, split by participants with and without a history of depression.

###### CAG Repeat Length

Residuals deviated from normality (W = 0.961, *p* = 0.005). The Q-Q plot showed some deviation, particularly at the extremes The box plot showed no evidence of heteroscedasticity with consistent variance. See eFig. 2.

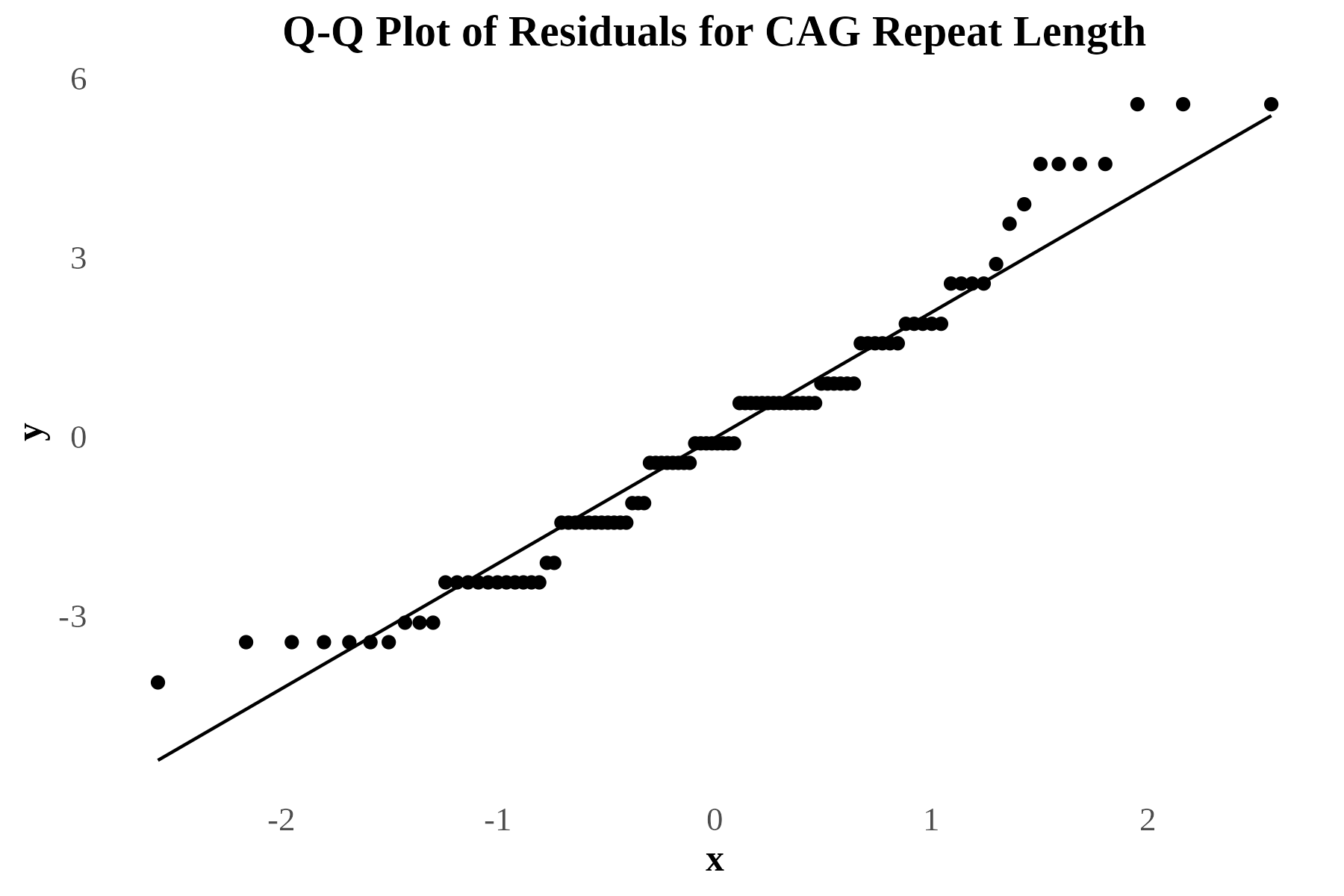

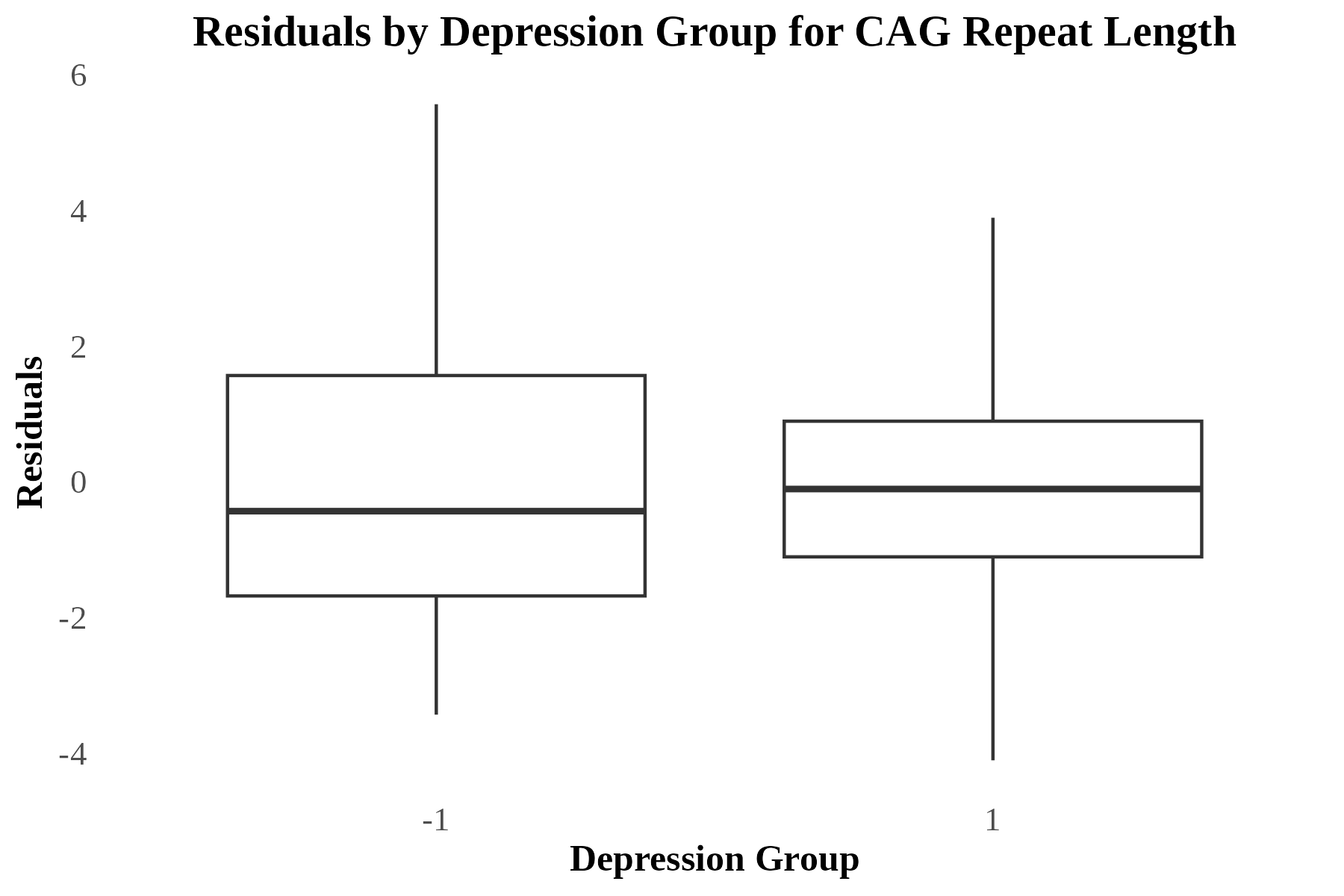

eFig. 2 Q-Q plot and boxplot of Residuals for Cytosine, Adenine, Guanine (CAG) Repeat Length. The left plot shows the results of the Q-Q plot of residuals for participant CAG repeat length. The right plot shows the results of the box plots of residuals for CAG repeat length, split by participants with and without a history of depression.

###### CAP Score

Residuals for were normally distributed (W = 0.978, *p* = 0.035), as supported by visual inspection of the Q-Q plot. The box plot showed no evidence of heteroscedasticity. See eFig. 3.

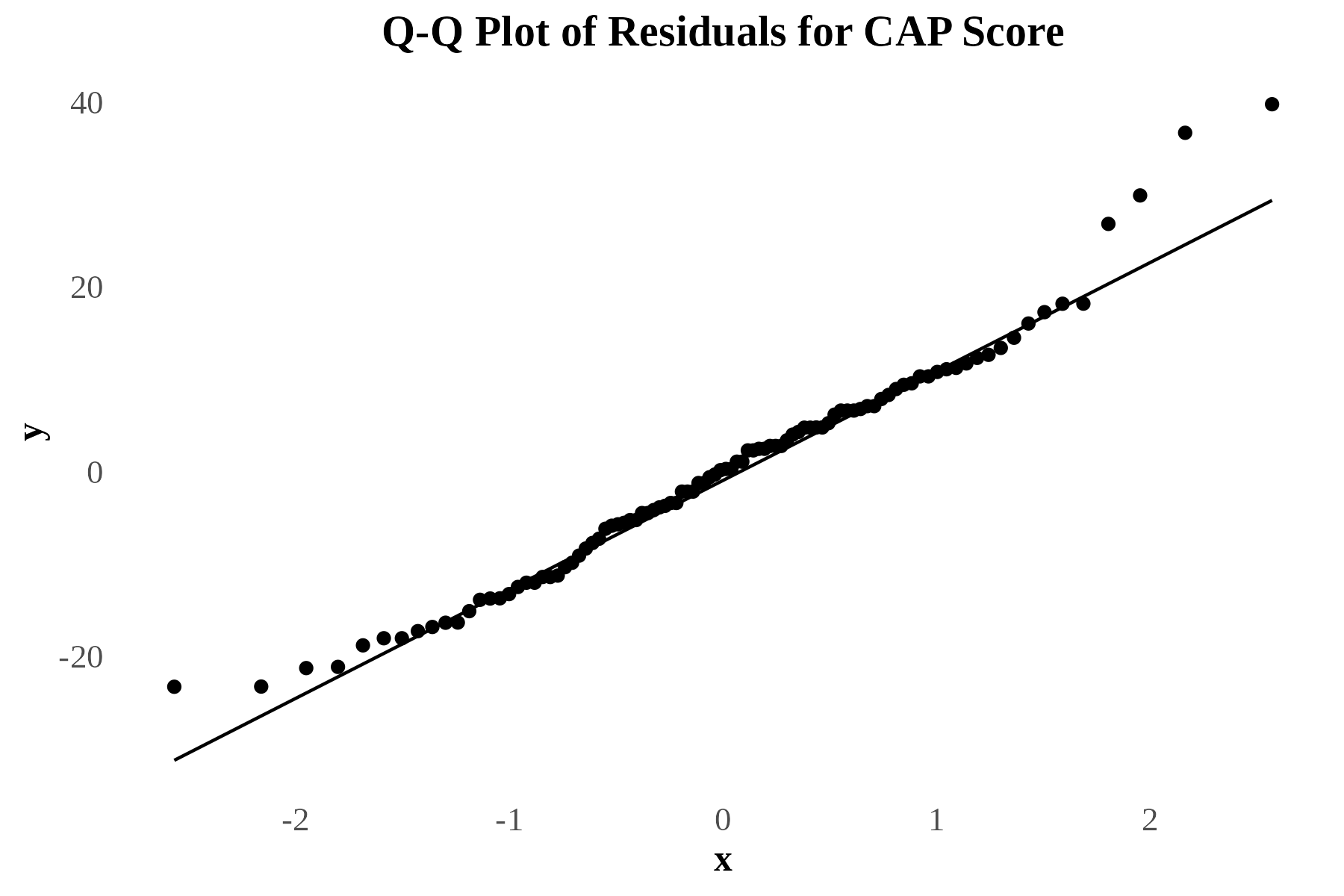

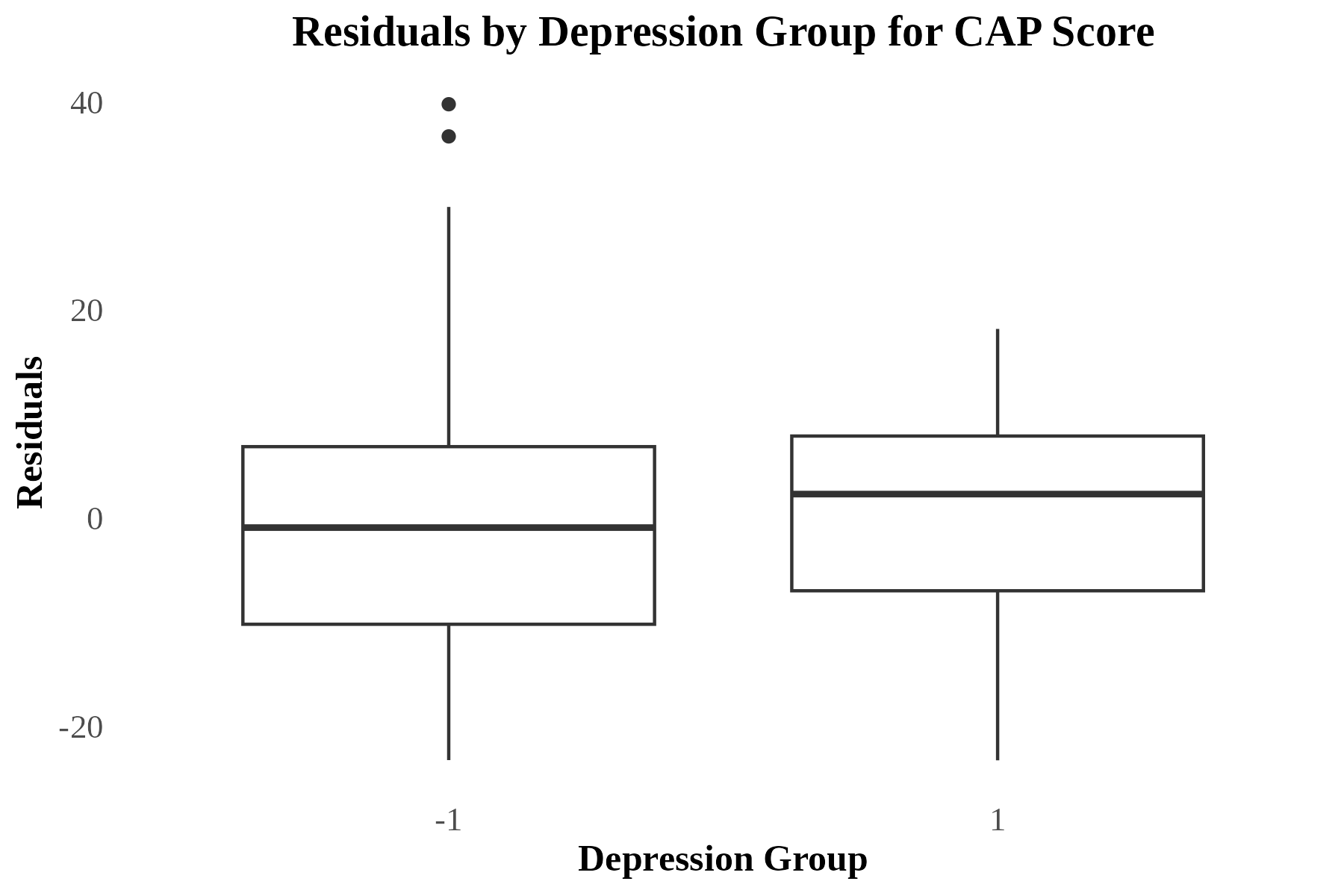

eFig. 3 Q-Q plot and boxplot of Residuals for CAG-Age Product (CAP) score. The left plot shows the results of the Q-Q plot of residuals for participant CAP Score. The right plot shows the results of the box plots of residuals for CAP Score, split by participants with and without a history of depression.

###### Disease Burden Score

Residuals were normally distributed (W = 0.992, *p* = 0.825), as supported by visual inspection of the Q-Q plot. The box plot showed no evidence of heteroscedasticity. See eFig. 4.

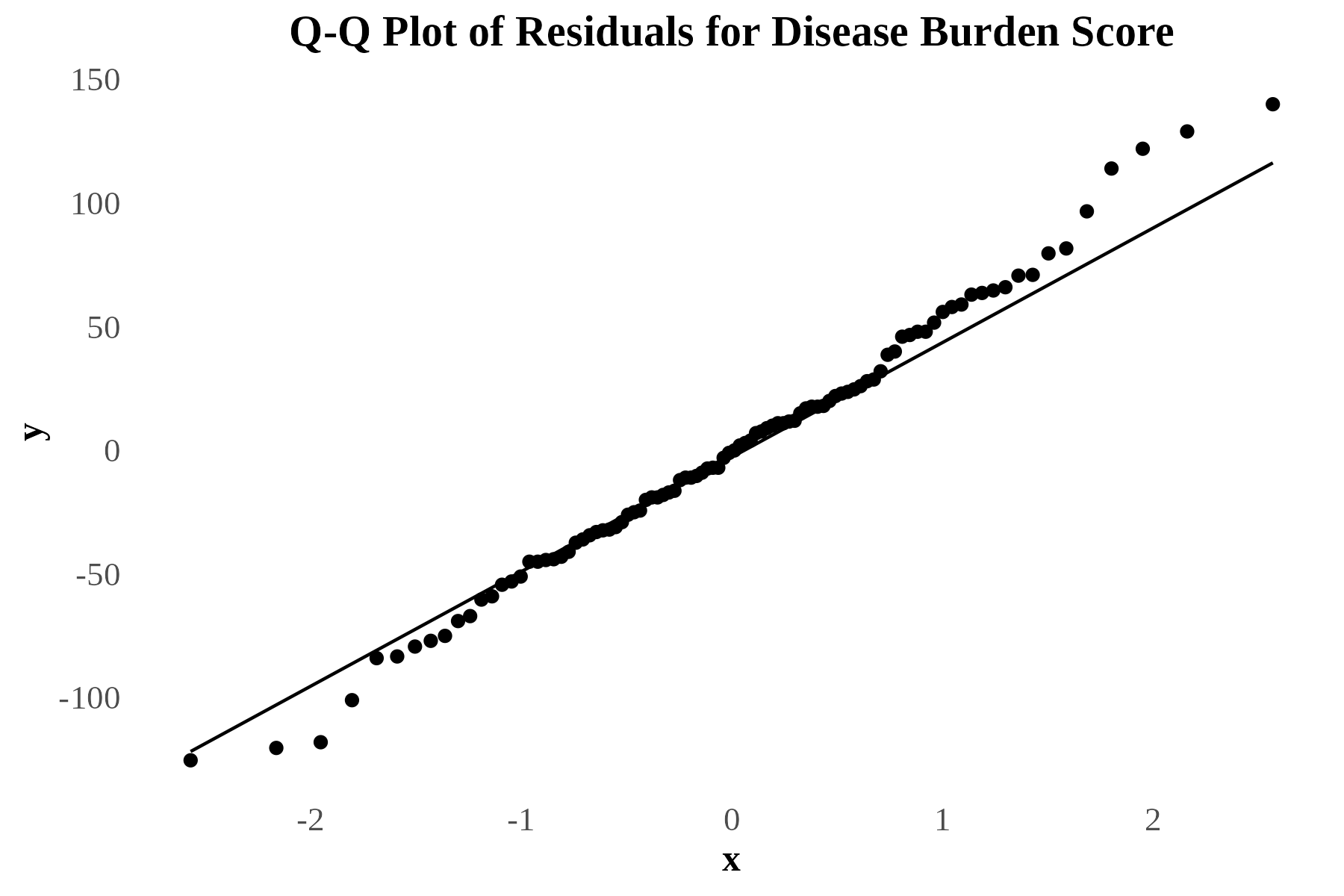

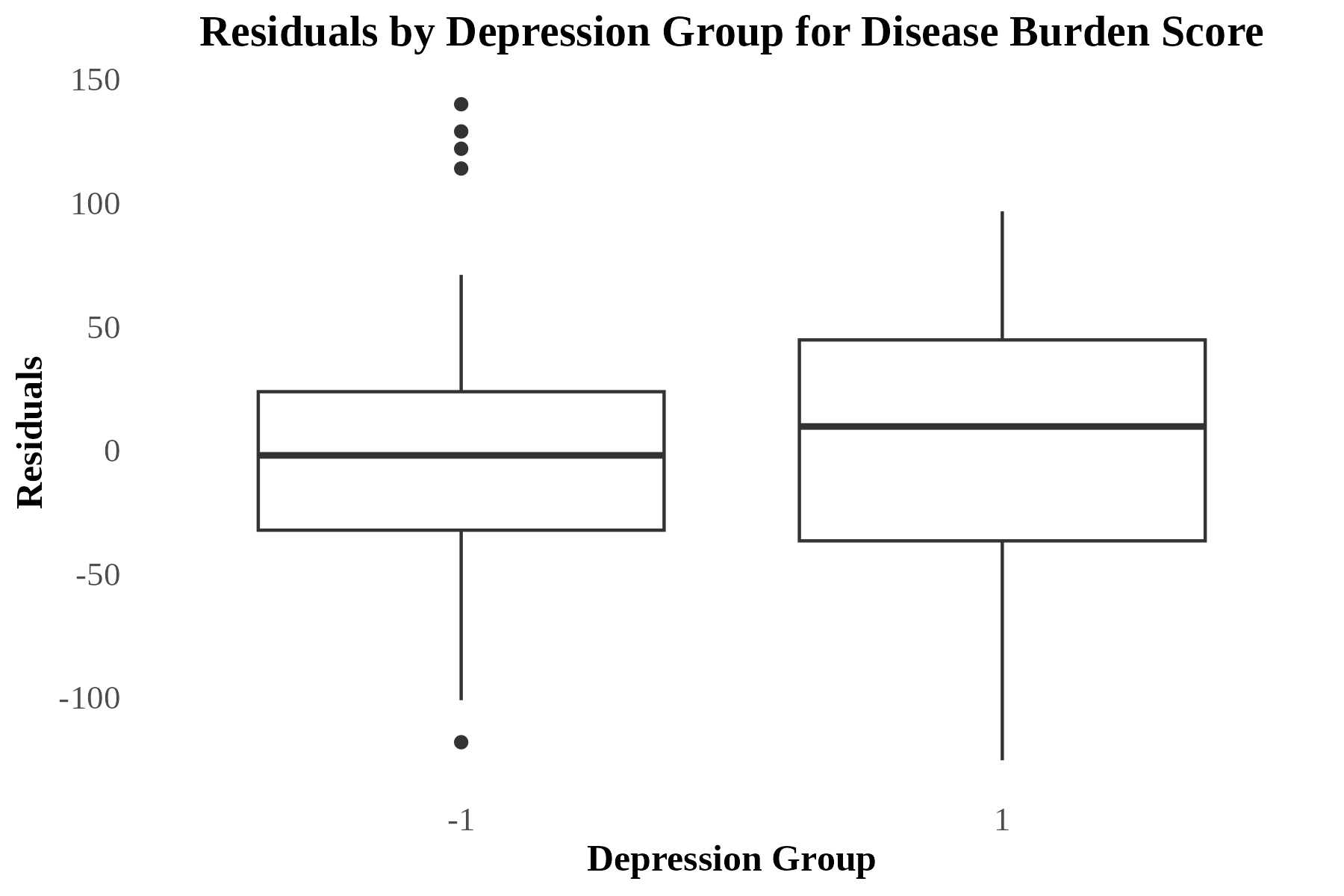

eFig. 4 Q-Q plot and boxplot of Residuals for Disease Burden Score (DBS). The left plot shows the results of the Q-Q plot of residuals for participant DBS. The right plot shows the results of the box plots of residuals for DBS, split by participants with and without a history of depression.

###### Unified Huntington’s Disease Rating Scale (UHDRS) Total Motor Score (TMS)

Residuals deviated from normality (W = 0.896, *p* < 0.001) and the Q-Q plot showed deviations at the tails. T The box plot showed no evidence of heteroscedasticity with consistent variance. See eFig. 5.

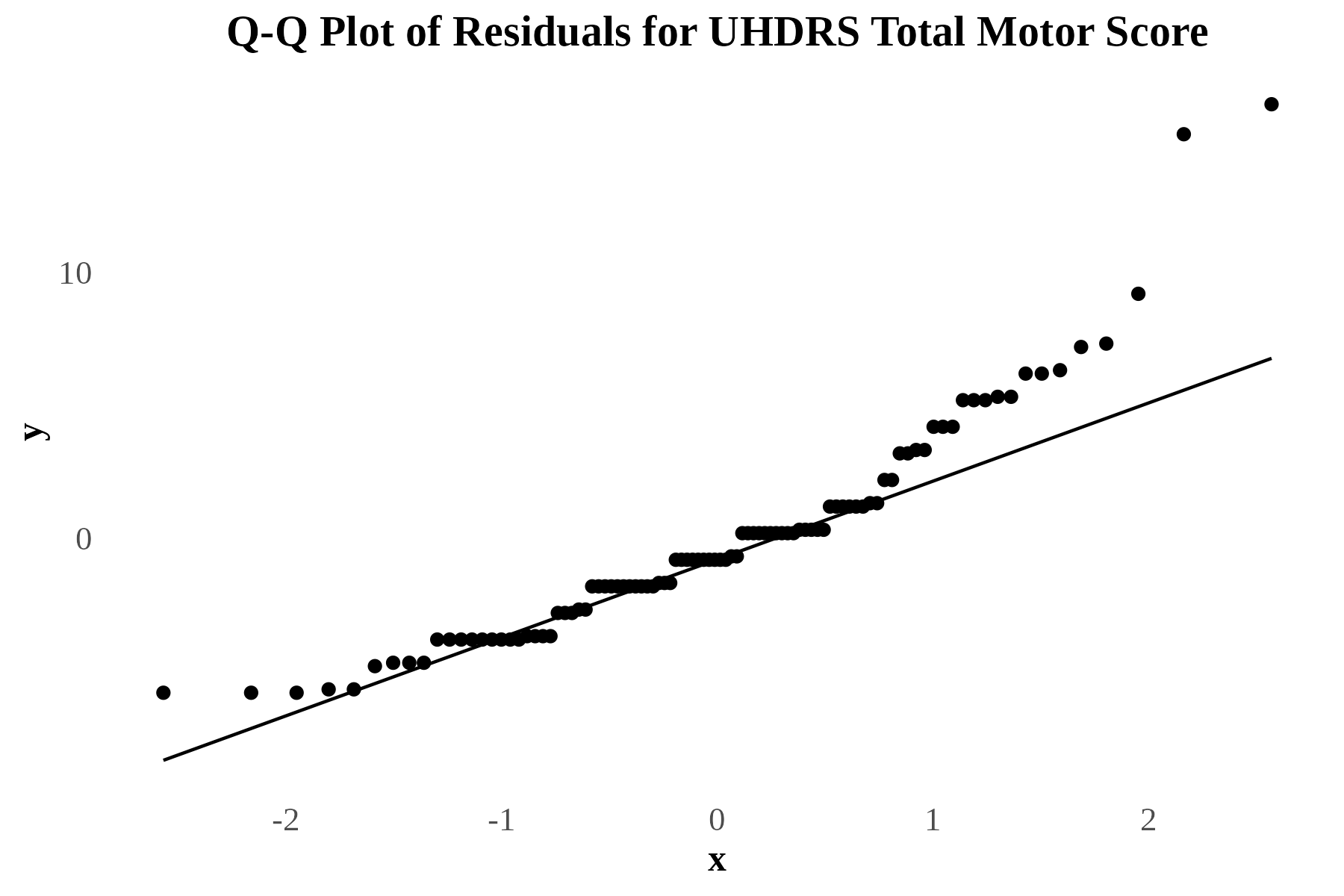

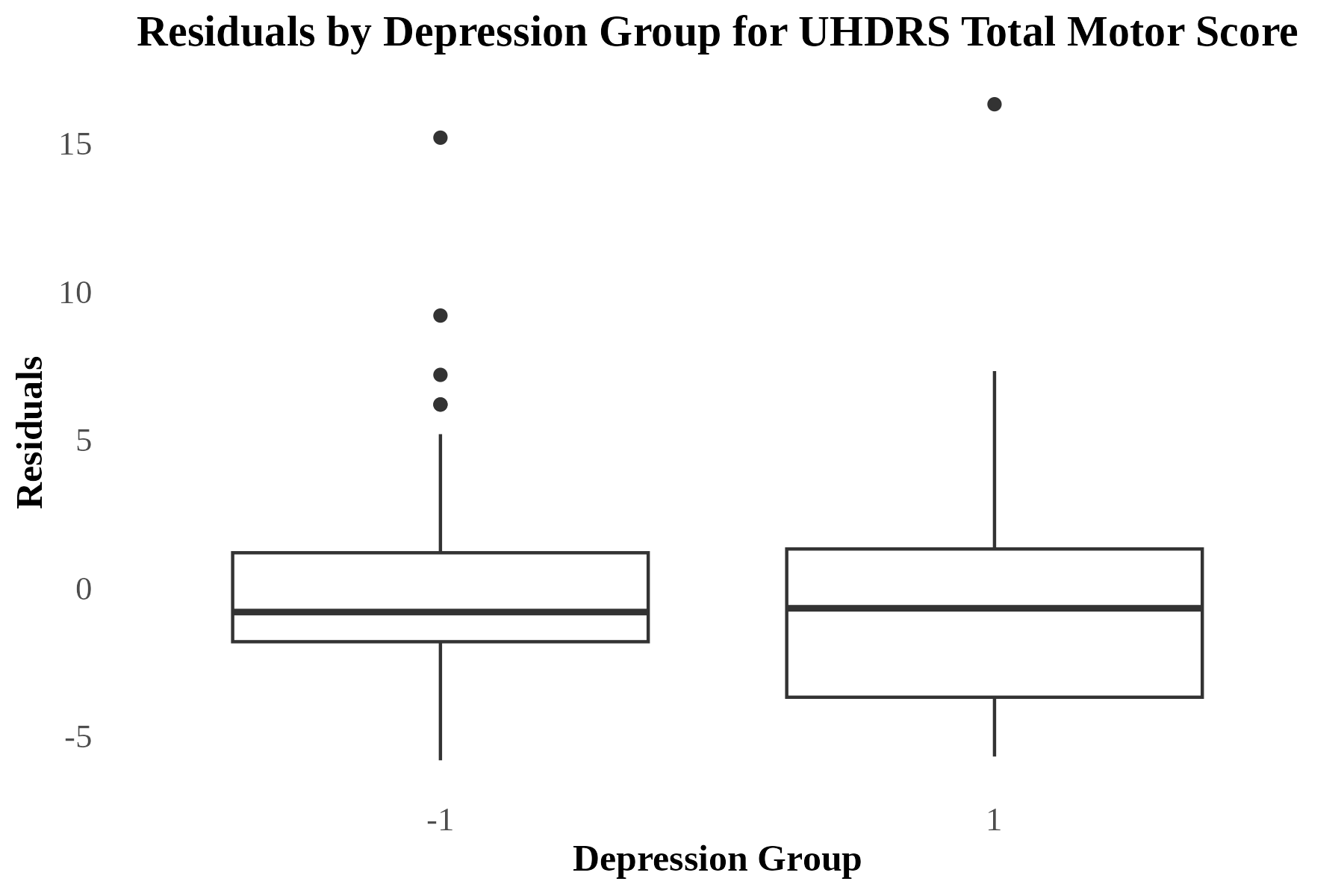

eFig. 5 Q-Q plot and boxplot of Residuals for Unified Huntington’s Disease Rating Scale (UHDRS) Total Motor Score (TMS). The left plot shows the results of the Q-Q plot of residuals for participant UHDRS TMS. The right plot shows the results of the box plots of residuals for UHDRS TMS, split by participants with and without a history of depression.

###### Beck Depression Inventory, 2nd Edition (BDI-II) Score

Residuals deviated from normality (W = 0.815, *p* < 0.001) and the Q-Q plot showed residuals deviating at the tails. The box plot showed a slightly wider spread in the depression group, indicating possible heteroscedasticity. See eFig. 6.

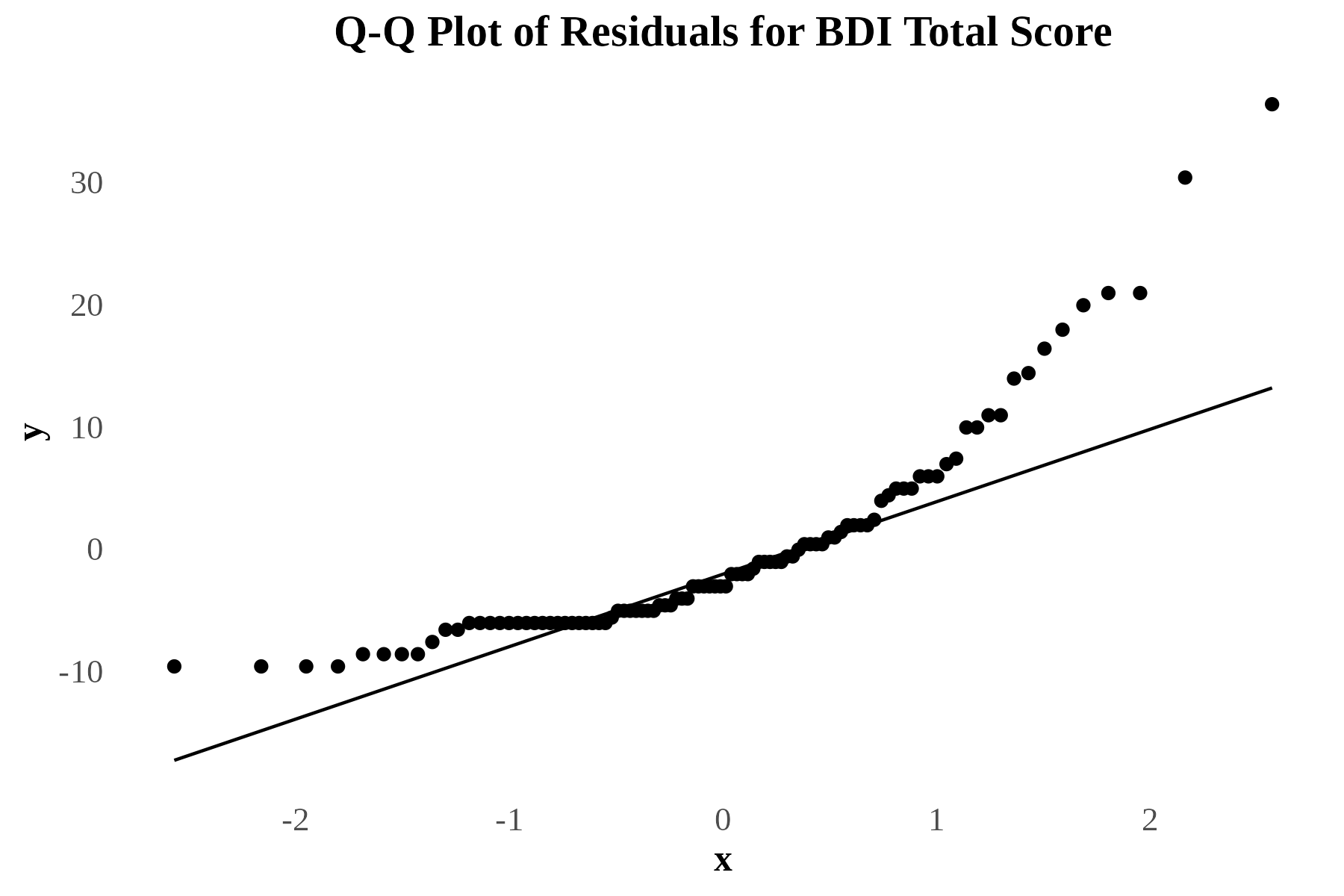

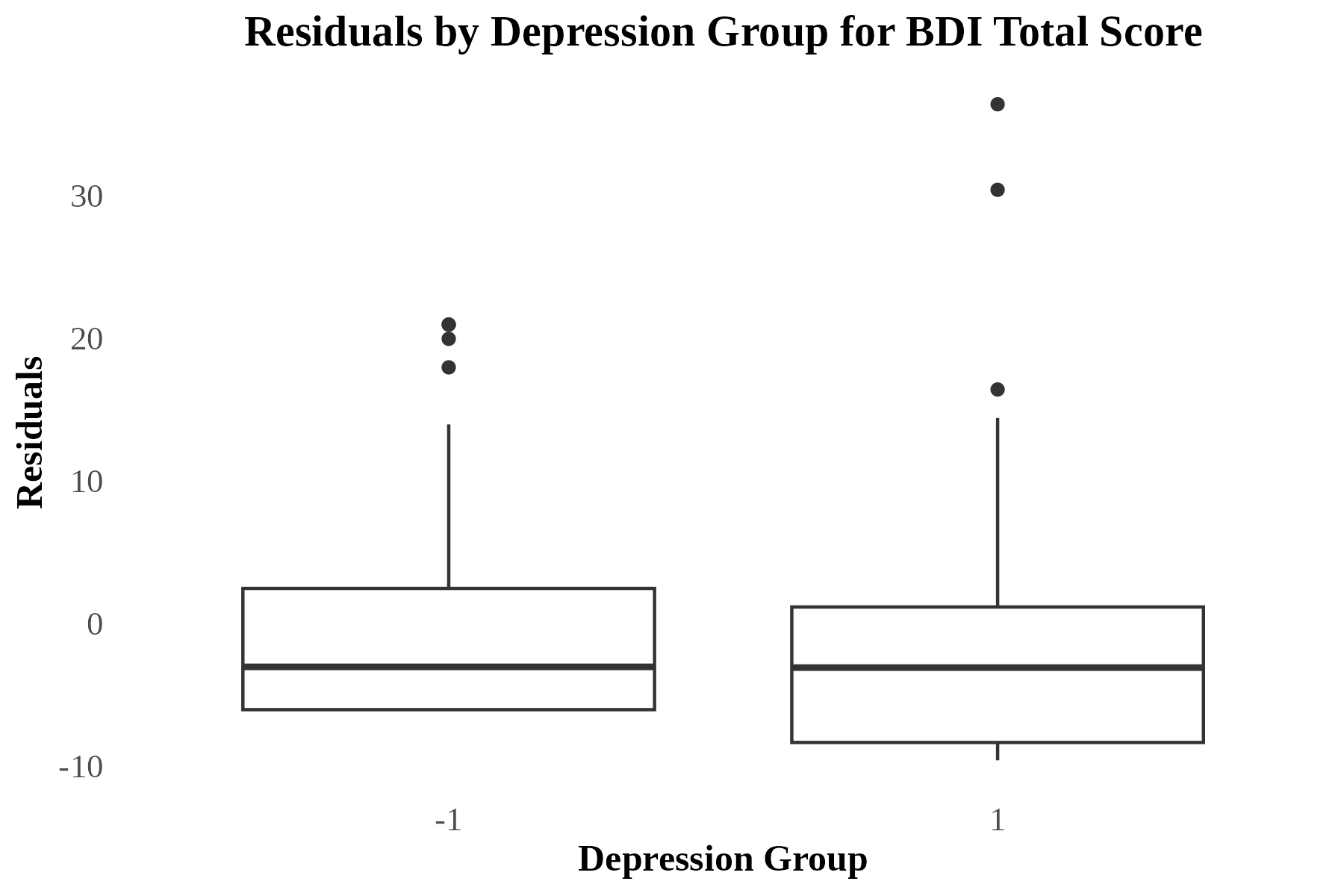

eFig. 6 Q-Q plot and boxplot of Residuals for Beck Depression Inventory, 2nd Edition (BDI-II) Score. The left plot shows the results of the Q-Q plot of residuals for participant BDI-II score. The right plot shows the results of the box plots of residuals for BDI-II score, split by participants with and without a history of depression.

###### Hospital Anxiety and Depression Scale, Depression Subscale (HADS-D) Score

Residuals deviated from normality (W = 0.796, *p* < 0.001) and the Q-Q plot showed residuals deviating at the tails. The box plot showed some outliers that contributed to non-normality but no evidence of heteroscedasticity. See eFig. 7.

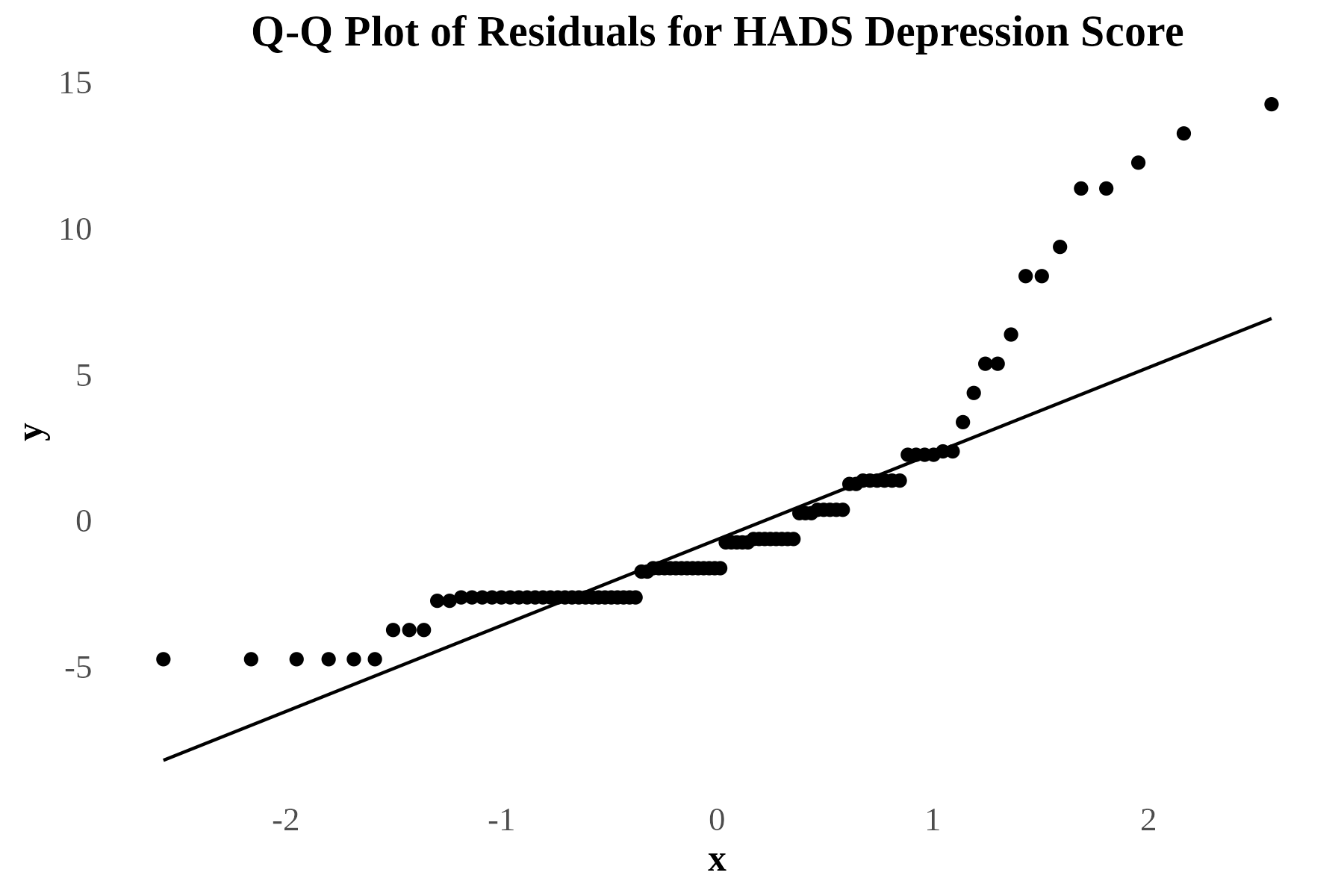

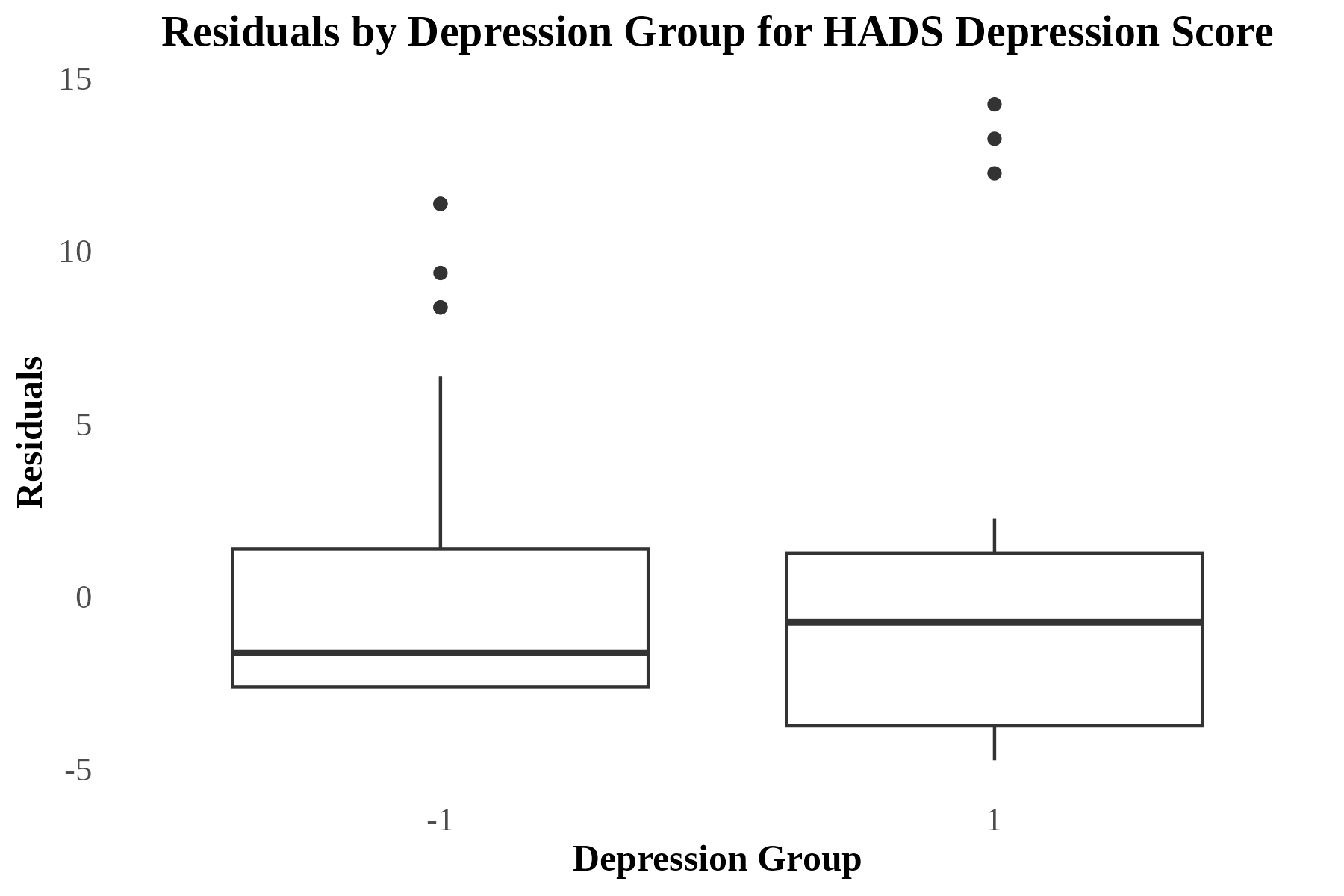

eFig. 7 Q-Q plot and boxplot of Residuals for Hospital Anxiety and Depression Scale, Depression Subscale (HADS-D) Score. The left plot shows the results of the Q-Q plot of residuals for participant HADS-D score. The right plot shows the results of the box plots of residuals for HADS-D score, split by participants with and without a history of depression.

#### Principal Component Analysis of Scanner Effects

Given that two different scanner systems were used, framewise displacement output from MRIQC v22.0.6(6) output were analysed to assess if there were significant differences in the scanner output. Visual inspection (see eFig. 8) and both Kolmogorov–Smirnov test and t-test were undertaken to assess for significant group differences. See eFig. 8.

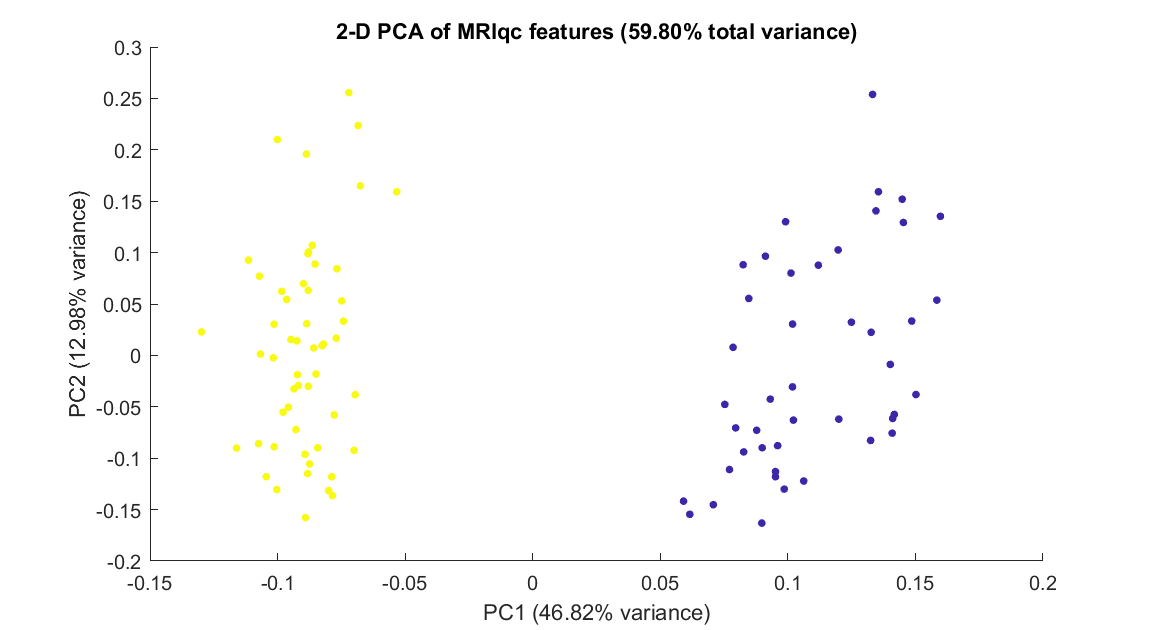

eFig. 8 Principal Component Analysis of Scanner Effects. Principal component analysis of MRI quality control metrics. The first two principal components are plotted, with data points colored by scanner type (blue = Siemens, yellow = Philips).

Framewise displacement differed significantly between scanner types, with Philips scanners showing higher mean FD than Siemens scanners (M = 0.224 vs. 0.177 mm; ***t***(96) = -2.41, ***p*** = .018; Kolmogorov-Smirnov ***D*** = 0.304, ***p*** = .017). The first principal component, explaining 46.82% of variance in MRI quality metrics, was included as a covariate in subsequent analyses to control for scanner-related quality differences.

#### Accuracy of DCM Model Estimation for HDGECs

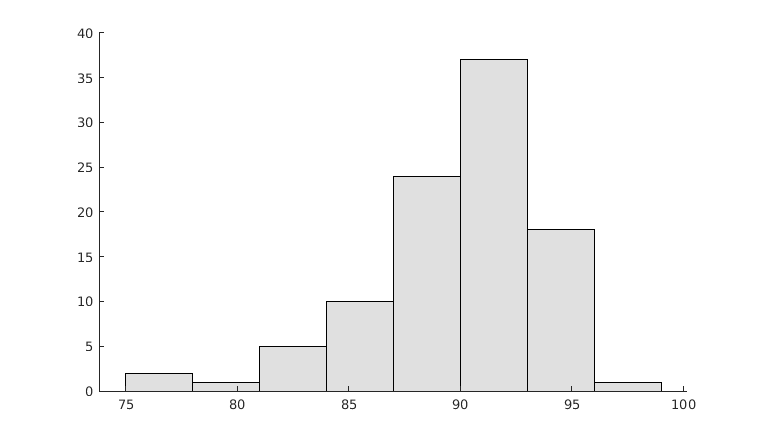

eFig. 9 Accuracy of DCM Model Estimation Across all Participants. The x-axis shows the percentage variance-explained by DCM model estimation for each participant, while the y-axis counts the number of participants with that percentage explained.

#### Region Effective Connectivity Changes Between Participants with a History of Depression and Without a History of Depression

eTable 3 summarises effective connectivity changes of the DMN and striatum for HDGECs associated with diagnosed depression history or HDGECs without depression. All reported connections met the significance criterion (free energy ≥ .95).

We investigated whether DMN and striatal effective connectivity changes underpin depression for HDGECs. We hypothesized increased effective connectivity from MPFC to PCC and hippocampus for participants with a history of depression compared to those without. We further hypothesized decreased effective connectivity from the MPFC to putamen and caudate for participants with a history of depression compared to those without, reflecting the early and severe degeneration of these regions. We expected to see a positive association between current depressive symptoms and increased connectivity from anterior to posterior DMN.

eTable 3 Between Region Effective Connectivity Changes Between Participants with a History of Depression and Those Without a History of Depression

| From → To | Valence | Posterior expectation | Credible interval |
| --- | --- | --- | --- |
| CAU_L_ → CAU_R_ | Excitatory | 0.07 | [0.01, 0.12] |
| CAU_L_ → PU_L_ | Inhibitory | 0.08 | [0.04, 0.13] |
| CAU_R_ → CAU_R_ | Inhibitory | 0.12 | [0.04, 0.20] |
| CAU_R_ → HPC_L_ | Inhibitory | 0.07 | [0.02, 0.12] |
| CAU_R_ → PCC | Inhibitory | 0.15 | [0.06, 0.23] |
| CAU_R_ → MPFC | Inhibitory | 0.10 | [0.03, 0.17] |
| PU_L_ → PCC | Inhibitory | 0.15 | [0.06, 0.24] |
| PU_L_ → MPFC | Inhibitory | 0.12 | [0.03, 0.18] |
| PU_R_ → CAU_L_ | Inhibitory | -0.14 | [-0.20, -0.08] |
| PU_R_ → PU_R_ | Inhibitory | 0.14 | [0.05, 0.22] |
| PU_R_ → MPFC | Inhibitory | 0.16 | [0.08, 0.23] |
| HPC_L_ → HPC_L_ | Inhibitory | -0.09 | [-0.17, -0.01] |
| HPC_L_ → MPFC | Inhibitory | 0.168 | [0.095, 0.241] |
| HPC_R_ → CAU_L_ | Inhibitory | -0.076 | [-0.132, -0.020] |
| HPC_R_ → PU_L_ | Inhibitory | 0.096 | [0.051, 0.142] |
| HPC_R_ → HPC_L_ | Inhibitory | -0.078 | [-0.135, -0.022] |
| HPC_R_ → PCC | Inhibitory | 0.108 | [0.030, 0.185] |
| PCC → HPC_L_ | Inhibitory | -0.054 | [-0.091, -0.017] |
| PCC → HPC_R_ | Inhibitory | -0.083 | [-0.117, -0.050] |
| MPFC → CAU_R_ | Excitatory | -0.054 | [-0.090, -0.019] |
| MPFC → HPC_R_ | Excitatory | 0.061 | [0.028, 0.094] |

**Note.** → = direction of connection; MPFC = Medial Prefrontal Cortex; PCC = Posterior Cingulate Cortex; HPC_L_ = Left Hippocampus; HPC_R_ = Right Hippocampus; CAU_L_ = Left Caudate; PU_L_ = Left Putamen; PU_R_ = Right Putamen.

#### Medication Sub-analyses Between HDGECs with a History of Depression and Those Without

The following sub-analysis replicated the effective connectivity changes between HDGECs with a history of depression compared to those without, but controlled for medication classes, resulting in eight binary covariates: antipsychotic use, benzodiazepine use, SSRI use, non-SSRI use, sex, and scanner effects.

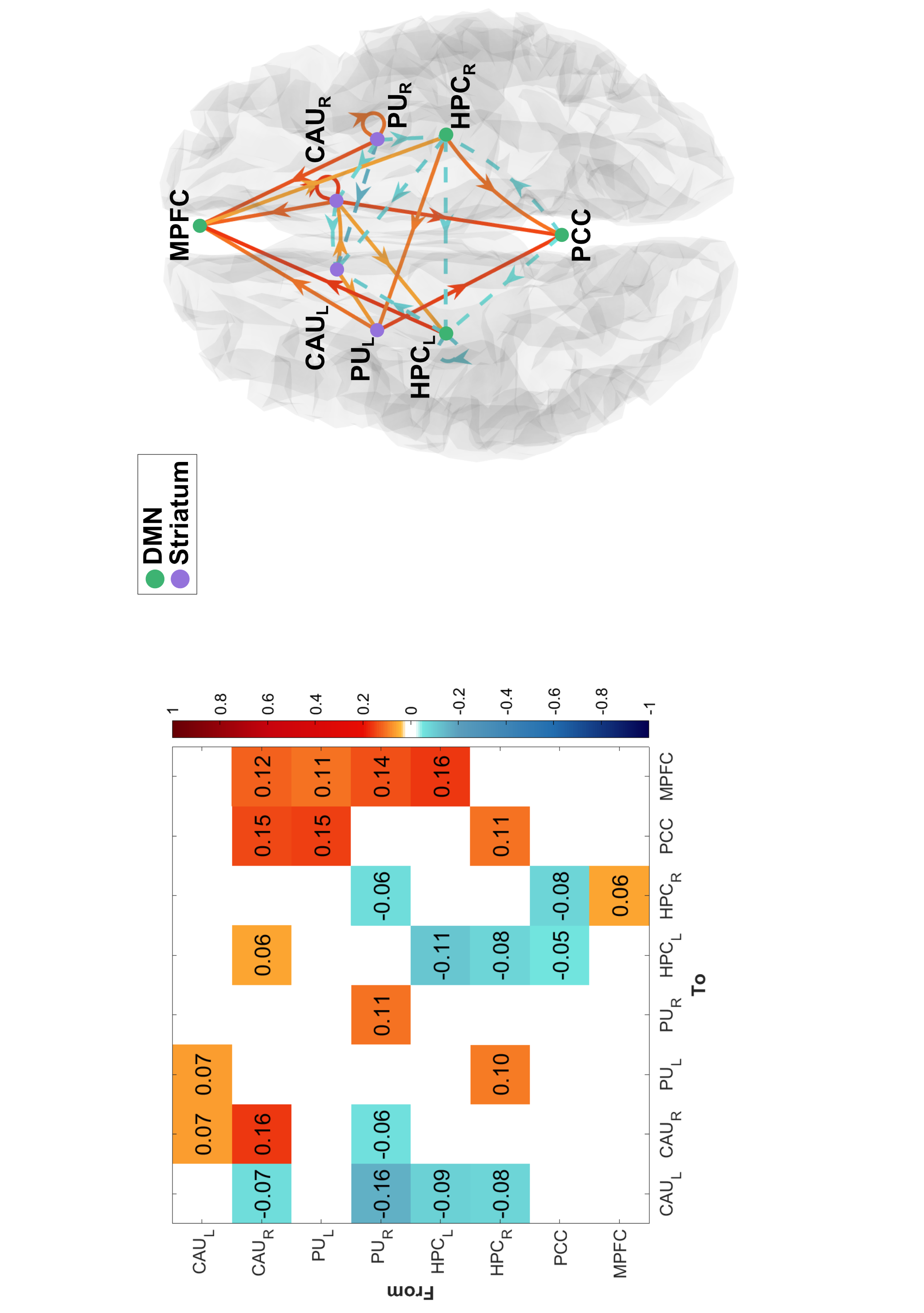

eFig. 10 Difference in Mean Effective Connectivity Between HDGECs with a History of Depression Versus Without, Modelling Separated Influence of Medications. (L) Left panel shows the connectivity matrix for depression history, with increased connectivity in yellow-red and decreased connectivity in aqua-navy. (R) Effective connections that reached significance are shown in the dorsal plane. Nodes in purple represent the bilateral caudate and putamen (striatum), while those in green represent the medial prefrontal cortex, posterior cingulate cortex, and bilateral hippocampi (default mode network). The arrows show the directed influence of one region on another, including self-connections. Solid lines represent increased connectivity while dashes represent decreased connectivity. MPFC = Medial Prefrontal Cortex; PCC = Posterior Cingulate Cortex; HPC_L_ = Left Hippocampus; HPC_R_ = Right Hippocampus; CAU_L_ = Left Caudate; CAU_R_ = Left Caudate; PU_L_ = Left Putamen; PU_R_ = Right Putamen.

The following sub-analysis removed all participants in both groups that were taking mood medications, resulting in 77 participants: 18 HDGECs with a history of depression and 59 HDGECs without a history of depression.

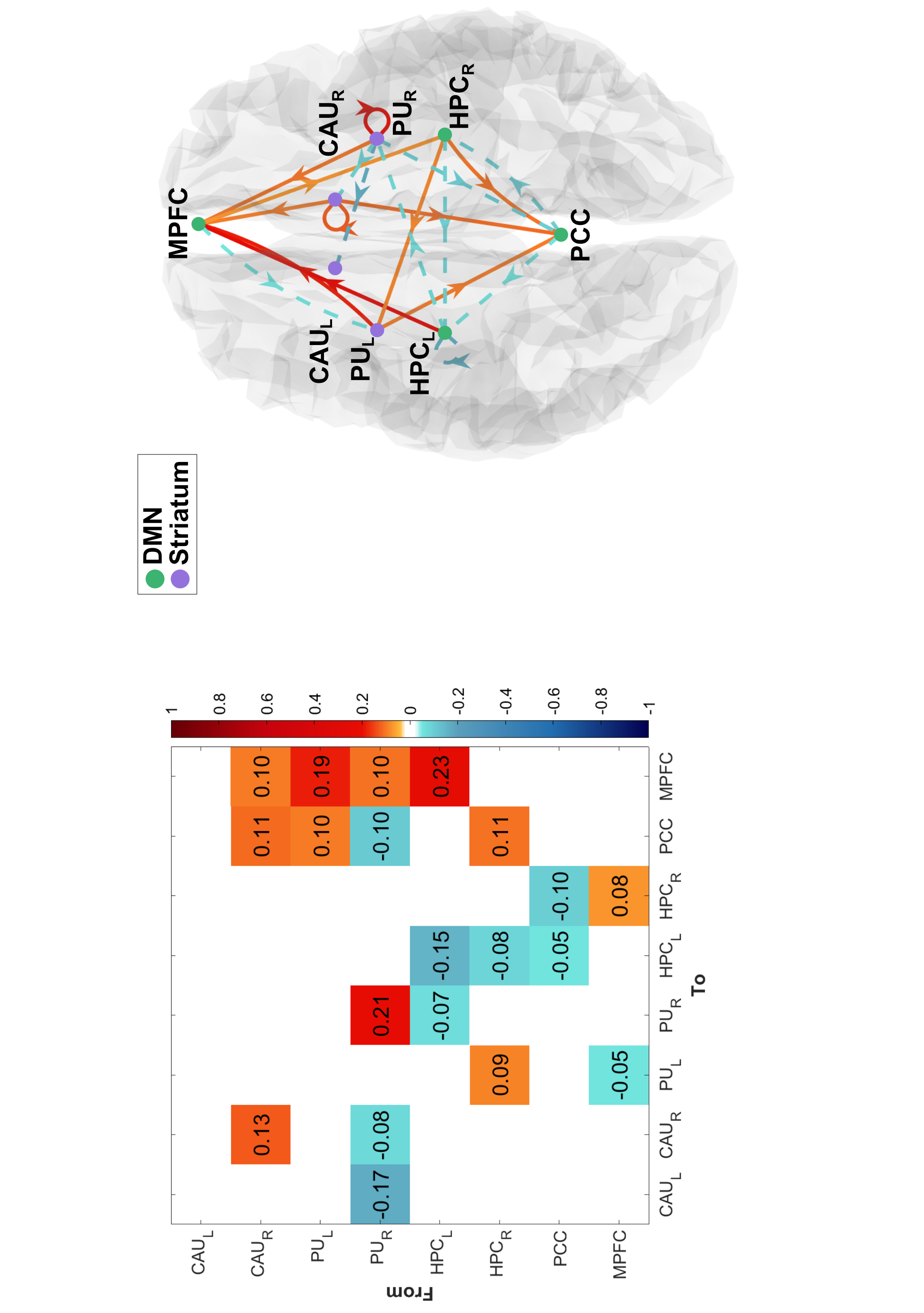

Difference in Mean Effective Connectivity Between HDGECs with a History of Depression Versus Without, for Participants not on Mood Medications. (L) Left panel shows the connectivity matrix for depression history, with increased connectivity in yellow-red and decreased connectivity in aqua-navy. (R) Effective connections that reached significance are shown in the dorsal plane. Nodes in purple represent the bilateral caudate and putamen (striatum), while those in green represent the medial prefrontal cortex, posterior cingulate cortex, and bilateral hippocampi (default mode network). The arrows show the directed influence of one region on another, including self-connections. Solid lines represent increased connectivity while dashes represent decreased connectivity. MPFC = Medial Prefrontal Cortex; PCC = Posterior Cingulate Cortex; HPC_L_ = Left Hippocampus; HPC_R_ = Right Hippocampus; CAU_L_ = Left Caudate; CAU_R_ = Left Caudate; PU_L_ = Left Putamen; PU_R_ = Right Putamen.

Regardless of how we controlled mood medication use, results were similar, supporting that a parsimonious model of one covariate sufficiently captured the variance in the BOLD signal accounted for by mood medication use. This also suggests that the modelling procedure of DCM, which discriminates neurovascular signals from neural ones, also parcellated the neurovascular impacts of mood medication sufficiently.

#### DMN and Striatal Connections as Brain Connectivity Associates of Depressive Symptoms

Leave-one-out cross-validation was performed using significant DMN connections from the group difference analysis, and analyses were completed on each individual connection, within DMN or striatum, DMN-to-striatum, striatum-to-DMN, and both PCC and left putamen, due to previous findings as potential disease hubs in HDGECs. Categories that had could classify the left-out participant are detailed in eTable 4.

eTable 4 Effective Connectivity Groups for Leave-one-out Cross-validation

| Connection Category | Effective Connections |
| --- | --- |
| Within Default Mode Network | HPC_L_ → HPC_L_, HPC_L_ → MPFC, HPC_R_ → HPC_L_, HPC_R_ → PCC, PCC → HPC_L_, PCC → HPC_R_, MPFC → HPC_R_ |
| From PCC | PCC → HPC_L_, PCC → HPC_R__ |

**Note.** → = direction of connection; ; HPC_L_ = Left Hippocampus; HPC_R_ = Right Hippocampus; MPFC = Medial Prefrontal Cortex; PCC = Posterior Cingulate Cortex.

We used the effective connectivity patterns to predict the left-out participant’s depression history status, and whether they met the clinically elevated cut-off for the HADS-D and BDI-II. Connections exceeding the significance criterion and significant correlations are reported in eTable 4.

eTable 5 DMN and Striatal Connections as Associates of Depression History and Current Depressive Symptoms

| From → To | Depression History | | | HADS-D | | | BDI-II | | |
| --- | --- | --- | --- | --- | --- | --- | --- | --- | --- |
|  | t(96) | p | r | t(96) | p | r | t(96) | p | r |
| PU_R_ → CAU_L_ | 2.45 | .008 | 0.243 | — | — | — | 2.56 | .006 | 0.253 |
| PU_R_ → MPFC | 1.88 | .032 | 0.187 | — | — | — | — | — | — |
| PCC → HPC_R_ | 2.07 | 0.02 | 0.207 | — | — | — | — | — | — |
| From PCC | 2.02 | .023 | 0.201 | — | — | — | — | — | — |
| Within DMN | 2.69 | .046 | 0.171 | — | — | — | — | — | — |

**Note.** Pearson’ s correlation coefficient is the association between observed values and predicted values for left-out participant. → = direction of connection; DMN = Default Mode Network. MPFC = Medial Prefrontal Cortex; PCC = Posterior Cingulate Cortex; HPC_L_ = Left Hippocampus; HPC_R_ = Right Hippocampus; CAU_L_ = Left Caudate; CAU_R_ = Left Caudate; PU_L_ = Left Putamen; PU_R_ = Right Putamen.

#### Mean Effective Connectivity within HDGECs

To understand the influence of the DMN in the models, we ran supplementary spDCM analyses looking at mean connectivity within each group: HDGECs with a history of depression and those without. Connections with free energy ≥ .95 are shown. eFig. 11 shows the connectivity matrices for both HDGECs with a history of depression and those without.

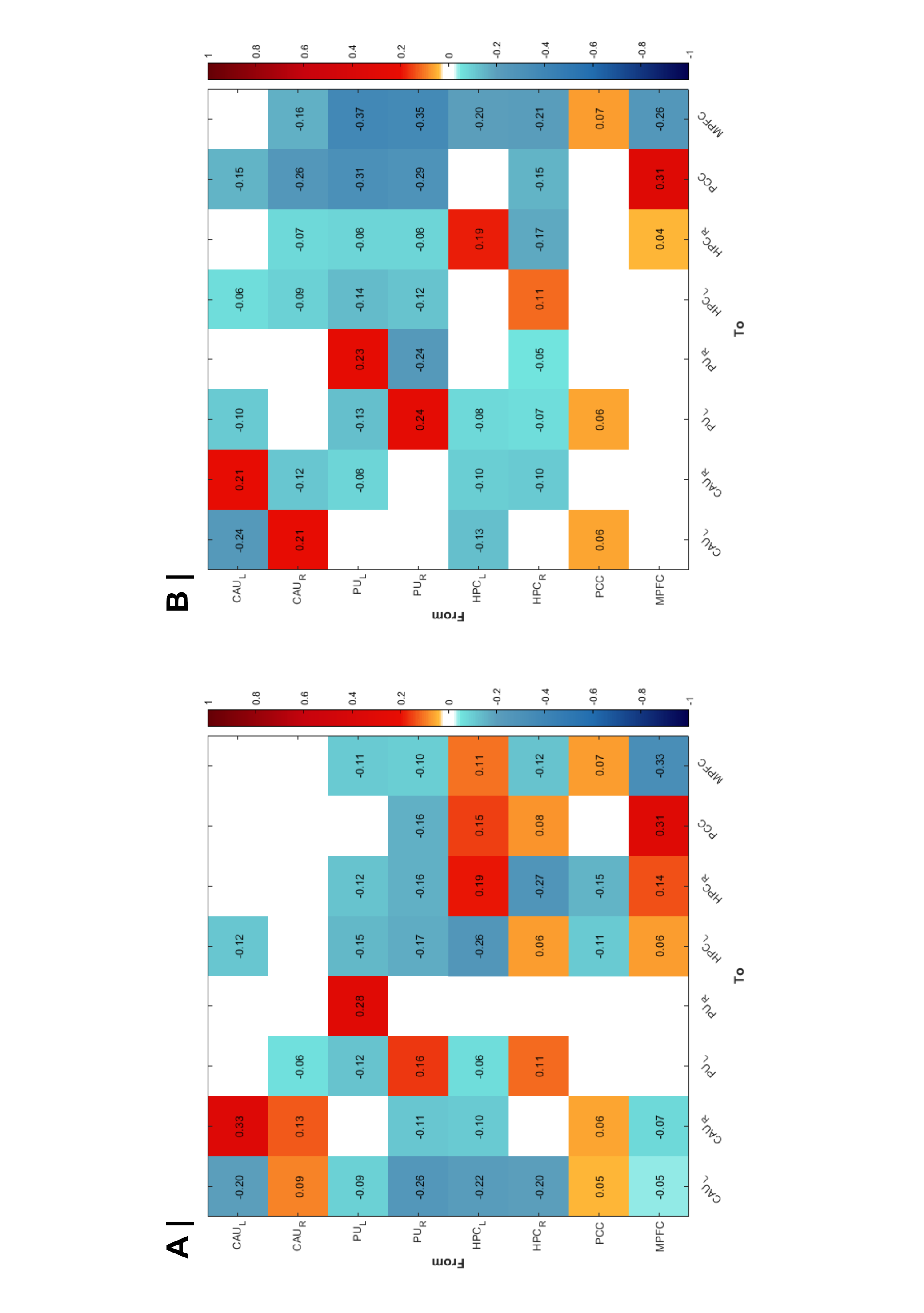

**eFig. 11 Mean Effective Connectivity of the Default Mode Network and Striatum for HDGECs with a History of Depression and Without. (A)** shows the mean connectivity within HDGECs with a history of depression. **(B)** shows the mean connectivity within HDGECs without a history of depression. Diagonal self-connections (intrinsic inhibitory connections) are displayed on a log scale to reflect their parameterization in DCM. Diagonal self-connections (intrinsic inhibitory parameters) are log-scaled in DCM and plotted on the same red/blue scale: red values reflect weaker self-inhibition (closer to zero decay), blue values reflect stronger self-inhibition (faster decay). Off-diagonal values follow the same convention, with red indicating excitatory influences and blue indicating inhibitory influences between regions.

We found excitatory influence of the MPFC on the PCC and increasing inhibitory self-connectivity of the MPFC for both groups. These analyses suggest that while MPFC has influence for both HDGECs with and without a history of depression, when we look at the differences these connections have cancelled each other out. This suggests that the MPFC is not the most salient change in connectivity for HDGECs with a history of depression compared to those with no history of depression.
